# Impact of close interpersonal contact on COVID-19 incidence: evidence from one year of mobile device data

**DOI:** 10.1101/2021.03.10.21253282

**Authors:** Forrest W. Crawford, Sydney A. Jones, Matthew Cartter, Samantha G. Dean, Joshua L. Warren, Zehang Richard Li, Jacqueline Barbieri, Jared Campbell, Patrick Kenney, Thomas Valleau, Olga Morozova

## Abstract

Close contact between people is the primary route for transmission of SARS-CoV-2, the virus that causes coronavirus disease 2019 (COVID-19). We sought to quantify interpersonal contact at the population-level by using anonymized mobile device geolocation data. We computed the frequency of contact (within six feet) between people in Connecticut during February 2020 – January 2021. Then we aggregated counts of contact events by area of residence to obtain an estimate of the total intensity of interpersonal contact experienced by residents of each town for each day. When incorporated into a susceptible-exposed-infective-removed (SEIR) model of COVID-19 transmission, the contact rate accurately predicted COVID-19 cases in Connecticut towns during the timespan. The pattern of contact rate in Connecticut explains the large initial wave of infections during March–April, the subsequent drop in cases during June–August, local outbreaks during August–September, broad statewide resurgence during September–December, and decline in January 2021. Contact rate data can help guide public health messaging campaigns to encourage social distancing and in the allocation of testing resources to detect or prevent emerging local outbreaks more quickly than traditional case investigation.

**One sentence summary:** Close interpersonal contact measured using mobile device location data explains dynamics of COVID-19 transmission in Connecticut during the first year of the pandemic.

## Introduction

Close contact between people is the primary route for transmission of the novel severe acute respiratory syndrome coronavirus 2 (SARS-CoV-2), the virus that causes coronavirus disease (COVID-19) [1]. Social distancing guidelines published by the United States (U.S.) Centers for Disease Control and Prevention (CDC) recommend that people stay at least six feet away from others to avoid transmission via direct contact or exposure to respiratory droplets [2]. Throughout the world, non-pharmaceutical interventions, including social distancing guidelines and stay-at-home orders, have been employed to encourage the physical separation of people and reduce the risk of COVID-19 transmission via close contact [3–6]. U.S. states with the lowest levels of self-reported social distancing behavior are experiencing most severe COVID-19 outbreaks [7].

While individual-level compliance with social distancing guidelines can be difficult to measure, researchers have proposed population-level mobility metrics based on mobile device geolocation data as a proxy measure for physical distancing and movement patterns during the COVID-19 pandemic [8–12]. Investigators have characterized geographic and temporal changes in mobility metrics following non-pharmaceutical interventions like social distancing guidelines and stay-at-home mandates during the COVID-19 pandemic [11, 13–22]. Researchers have also studied the association between mobility metrics and COVID-19 cases or other proxy measures of transmission [11, 23–34]. Most mobility metrics measure aggregated movement patterns of individual mobile devices: time spent away from home, distance traveled, or density of devices appearing in an area during a given time interval. CDC reports mobility metrics from Google, Safegraph, and Cuebiq [35]. Some mobility metrics measure spatial relationships among individual devices. Klein et al. [12] measure “colocation” events, in which reported locations of two devices lie within a roughly 60 square foot spatial grid cell. Couture et al. [16] compute a “device exposure index” that measures the colocation of devices within a sample of preselected venues like restaurants or retail establishments. Chang et al. [30] use colocation matrices from Facebook [36] that measure the probability that devices from different geographic areas appear in the same 600-meter square region for five minutes, aggregated by week. Morley et al. [37] use the “human encounters” metric from Unacast [38, 39] that measures the frequency of two devices being within 50 meters of each other for an hour or less. Finally, Cuebiq offers a contact index measuring when two or more devices are within 50 feet of each other within five minutes [40].

Existing mobility metrics might not capture simultaneous colocation of devices, do not measure contact within a two-meter distance associated with highest transmission risk (via direct contact or exposure to respiratory droplets), and might not take intrinsic mobile device spatial location error (horizontal uncertainty) into account. A better measure of contact events – the primary behavioral risk factor for transmission – could help explain historical patterns of transmission, assist policymakers in targeting interventions and messaging campaigns to encourage social distancing, guide public health response measures such as enhanced testing and contact tracing, and provide early warning to detect and prevent emerging outbreaks. By using highly detailed mobile device geolocation data and a novel probabilistic method for assessing close proximity, we sought to quantify total intensity of close interpersonal contact (within six feet) at the population-level (contact rate) and to use contact rate to explain patterns of COVID-19 incidence and predict emergence of new COVID-19 cases in the state of Connecticut, U.S. during February 1, 2020 – January 31, 2021.

## Setting: Connecticut

Connecticut (population 3.565 million), like other states in the northeastern U.S., experienced a strong initial wave of COVID-19 infections during March–April 2020 following outbreaks in the New York City area [41, 42]. On March 17, Connecticut Governor Ned Lamont closed schools [43–46], and issued a statewide “Stay Safe, Stay Home” mandate to take effect on March 23, 2020 [47]. Governor Lamont’s executive order recommended that nonessential businesses cease all in-person functions, closed in-person dining at restaurants, and cancelled all in-person community gatherings. The mandate excluded healthcare, food service, law enforcement, and other essential services.

As case counts declined, Connecticut followed a gradual reopening plan designed to resume economic activity while minimizing the risk of transmission via close contact between people. On May 20, the state entered Phase 1, permitting the following to open at 50% capacity with social distancing: hair salons and barbershops, outdoor zoos and museums, outdoor dining, outdoor recreation, retail shopping, university research, and offices, although work from home was strongly encouraged [48]. On June 17, Phase 2 began, permitting indoor religious services at 25% capacity and capped at 100 people, outdoor religious services capped at 150 people, and opening indoor dining, hair salons, personal service businesses, and libraries at 50% capacity [49]. A serology study to measure prevalence of SARS-CoV-2-specific IgG antibodies was conducted among adult Connecticut residents residing in non-congregate settings during June–July [50]. The study estimated a seroprevalence of 4.0% (90% confidence interval 2.0%–6.0%). Participants in the study reported their risk mitigation behaviors: 73% avoided public places, 75% avoided gatherings of families or friends, and 97% wore a mask at least some of the time. In July, Governor Lamont delayed the state’s planned summer move to Phase 3 – which would have loosened occupancy restrictions on bars and restaurants – because of surges in transmission occurring elsewhere in the U.S. [51].

Connecticut experienced low COVID-19 incidence and declining hospitalization during June–August, but in August a major outbreak occurred in Danbury, a town in the western part of the state [52]. During August–September, in-person education resumed at many colleges, universities, and primary/secondary schools in Connecticut. By mid-September, the state was facing a broad resurgence of COVID-19 transmission. On September 17, the Connecticut Department of Public Health reported that the number of new cases per week for the previous 4 weeks was 62% higher than the average number of new cases per week in July and early August [53]. These signs of resurgence were initially concentrated in southeastern Connecticut, where few COVID-19 cases were identified during the initial spring wave in March–April [54, 55].

Public health officials identified travel, social gatherings, workplaces, churches, universities, and recreational sports as contributing to transmission [56]. Nevertheless, on October 8, the state began Phase 3 reopening, permitting 50% capacity in houses of worship capped at 200 people, uncapped outdoor religious gatherings with social distancing, and opening indoor dining, hair salons, personal service businesses, and libraries at 75% capacity [49]. On November 6, as COVID-19 case counts continued to increase, Connecticut reverted to “Phase 2.1”, reducing indoor restaurant seating, indoor and outdoor event capacity, and placing caps on attendance [49]. Case counts increased through December and began to decline in January 2021. Overall, Connecticut residents complied with state guidelines and mandates to reduce close contact. In a survey of risk mitigation behaviors throughout the U.S., Lazer et al. [7] reported that Connecticut ranked 9th among U.S. states in self-reported social distancing during fall 2020, and 6th in self-reported mask wearing. But case counts indicate that Connecticut experienced widely varying temporal and geographic dynamics of COVID-19 incidence over the course of the pandemic.

## Computing contacts from anonymized mobile device geolocation data

We obtained anonymized mobile device geolocation data for a sample of devices in Connecticut from X-Mode. During May 1, 2020 through January 31, 2021, we observed a total of 788,842 unique (anonymized) device IDs, representing roughly 22% of the approximately 3.565 million residents of Connecticut (though some of those devices may have belonged to people residing elsewhere). An average of 141,617 unique devices were observed per day. For each week, an average of 80.5% of device IDs from the prior week were present in the data. Devices might not be present in the dataset if the user turns off the device or does not interact with applications that report location data. Using device geolocation records consisting of anonymized device IDs, GPS coordinates, date/time stamps, and GPS location error estimates (horizontal uncertainty), we calculated the location in which each device had the most location records and designated that area as the device’s primary dwell location (i.e., town of residence of device owner).

A contact event was computed by using a probabilistic algorithm that computes the likelihood of simultaneous 2-meter proximity between pairs of devices across geographic areas. For each device, we identify sets of records where devices were in spatial proximity to one another and stationary. A limitation of mobile device gelocation data is that it is not possible to precisely quantify the duration a device is stationary because device locations are collected asynchronously and irregularly over time. For each potential contact event, we compute the probability that the two device locations are within six feet by assuming that the reported device locations arise from a two-dimensional Gaussian probability distribution whose variance is computed by using the horizontal uncertainty measure, and correct the distance to account for the curvature of the earth. Figure S1 shows a schematic illustration of the contact event probability calculation.

**Figure 1:**
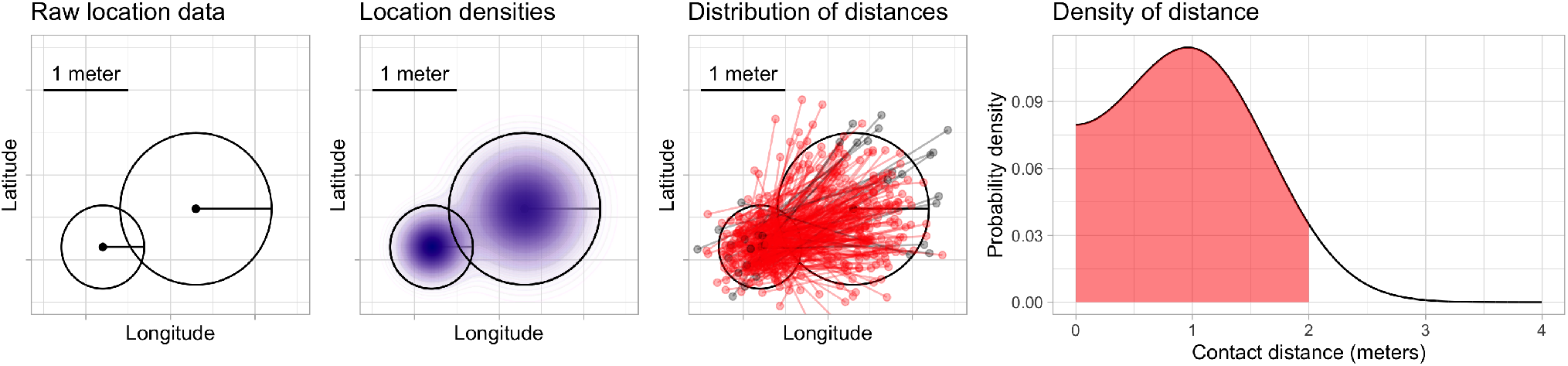
Schematic illustration of contact probability calculation. From left to right: raw locations, including horizontal uncertainty estimates, for two mobile devices are transformed into approximate location probability densities. The distribution of distances from points drawn randomly from these densities is computed. Sampled distances are shown here for illustrative purposes in red (when sampled device locations are within six feet apart) and gray (when sampled locations are more than six feet apart); in our implementation, the distribution of these distances is computed analytically. The shaded area under the density is the probability that the devices are within six feet.

We define the “contact rate” as the total number of contact events per day among observed devices at the town level; the contact rate is computed by summing daily contact probabilities for each device and assigning that sum to the device primary dwell location. A detailed description of the mobile device geolocation data, computation of the probability of contact, spatial aggregation of the contact probabilities to estimate contact rate, and coverage of mobile devices across Connecticut is given in the Supplementary Materials.

The mobile device geolocation and COVID-19 case data contain no individually identifying meta-data and were aggregated by day and town. This work was approved by the Yale University institutional review board. This work was also reviewed by CDC and was conducted consistently with applicable federal law and CDC policy ^1^.

## Statewide contact trends

Figure S2 shows the contact rate by town in Connecticut during February 1 – January 31, 2021. Maps show the weekly average of daily contact rate by town, where darker colors in maps indicate higher contact rate. The daily contact rate is shown in the plot below. The statewide contact rate dropped dramatically in March, about one week before Governor Lamont issued the statewide stay-at-home mandate on March 23 [47]. News of surging COVID-19 hospitalization and responses in the New York area [57, 58], closure of public schools, and anticipation of a possible stay-at-home order might have played a role in reducing contact before the mandate was announced. After staying low during most of April, the contact rate began to rise slowly throughout the state during June–August. Incidence of infection was likely much higher during the first wave than the second, but steadily increasing availability of SARS-CoV-2 testing yielded higher case counts in the second wave. An interactive web application for exploring the contact rate in Connecticut is available at https://forrestcrawford.shinyapps.io/ct_social_distancing. The Supplementary Materials describe the web application in detail.

**Figure 2:**
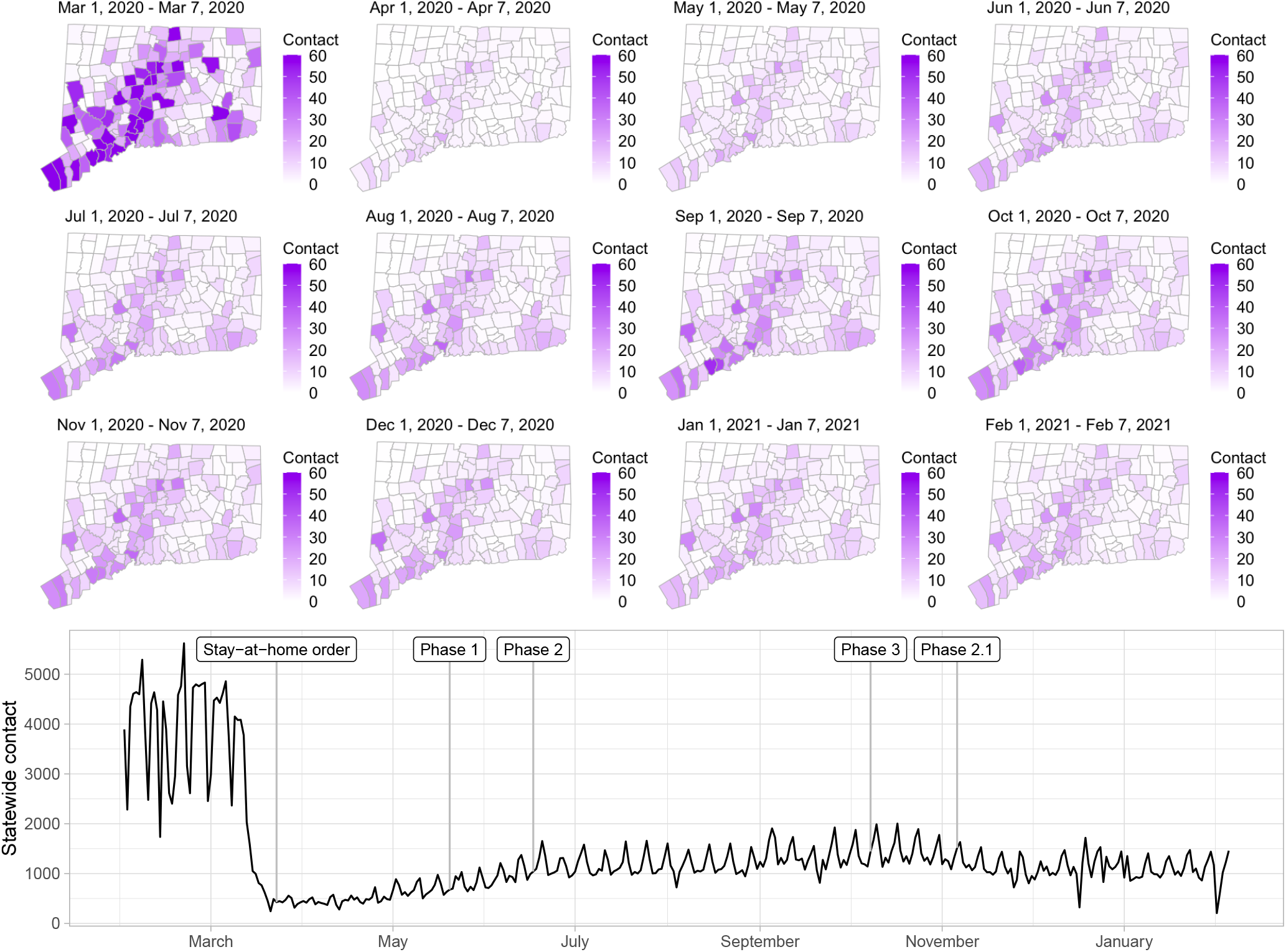
Estimated contact rate among mobile devices in our dataset in Connecticut from February 2020 to February 2021. At top, maps show the number of contacts in Connecticut’s 169 towns per day during weeks beginning on the first of each month. Darker colors indicate higher contact. At bottom, statewide contact shows the daily frequency of close contact within six feet between distinct devices in our dataset. Governor Ned Lamont’s stay-at-home order and reopening phases 1, 2, 3, and 2.1 indicated. The state reverted to the more restrictive “Phase 2.1” in response to rising case counts in November.

The Supplementary Material presents a comparison of the contact rate to mobility metrics from Google [59], Apple [60], Facebook [61], Descartes Labs [14, 62], and Cuebiq [40]. Most mobility metrics provided by these companies returned to values near the February/March baseline by the beginning of July. In contrast, the contact rate shown in Figure S2 shows that close interpersonal contact stayed low and rose slowly during June–August, 2020. Mobility metrics returned more quickly to the February 2020 baseline (or higher) compared to the contact rate and do not explain the low COVID-19 incidence achieved in Connecticut during June–August, 2020.

One explanation for the discrepancy between close contact and mobility metrics is that it is possible to travel far from home, to many distinct points of interest, or to many geographic areas, without coming into close contact with others. This might be what occurred in the summer of 2020: as Connecticut began its phased reopening plan, people resumed more normal patterns of away-from-home movement – work, shopping, or recreational activities – while maintaining social distancing. For this reason, when mobility metrics are used as proxy measures of close interpersonal contact, they might overstate the risk of disease transmission.

## Prediction of COVID-19 cases in Connecticut towns

To evaluate the contact rate as a predictor of COVID-19 burden in Connecticut, we use confirmed COVID-19 case data from non-congregate settings reported to the Connecticut Department of Public Health. We excluded cases among residents of long-term care facilities, managed residential communities (e.g., assisted living facilities), or correctional institutions. We aggregated non-congregate case data by day of sample collection, by town. We obtained town-level population estimates from the American Community Survey [63, 64].

We predict transmission of SARS-CoV-2 and COVID-19 cases in Connecticut towns using a continuous-time deterministic compartmental transmission model based on the susceptible-exposed-infective-removed (SEIR) process [65]. We accommodate geographical variation in transmission within Connecticut and estimated features of COVID-19 disease progression, hospitalization, and death. The model incorporates flexible time-varying case-finding rates at the town level. We incorporate the contact rate into the time-varying transmission risk by multiplying the standardized contact rate by the product of the baseline transmission rate and the estimated number of susceptible and infectious individuals in each town. We fit the model to statewide data, and produce model projections for each of Connecticut’s 169 towns using the town population size, time-varying contact rate, estimated initial infection fraction, and time-varying case-finding rate. The model is conceptually similar to other SEIR-type COVID-19 transmission models making use of mobility data, but incorporates much geographic variation in transmission rates [26, 66–74]. The model and calibration procedure are described in detail in [65] and in the Supplementary Materials.

Figure S3 shows contact rates, estimated SARS-CoV-2 infections, observed and estimated case counts, estimated cumulative incidence, as well as 95% uncertainty intervals for model estimates, for the five largest cities by population in Connecticut: Bridgeport, Hartford, New Haven, Stamford, and Waterbury. Contact rates in these towns largely mirror rates in the state as a whole. Model estimates track the pattern of case counts through the full course of the epidemic, including the dramatic reduction in transmission during June–August. In some towns, e.g. Stamford, case counts are under-estimated in model projections during the first wave during March–April 2020. In these cases, dynamics of SARS-CoV-2 infections may differ from the dynamics of case counts because the estimated case detection rate (via viral testing) varied dramatically over time and geography.

**Figure 3:**
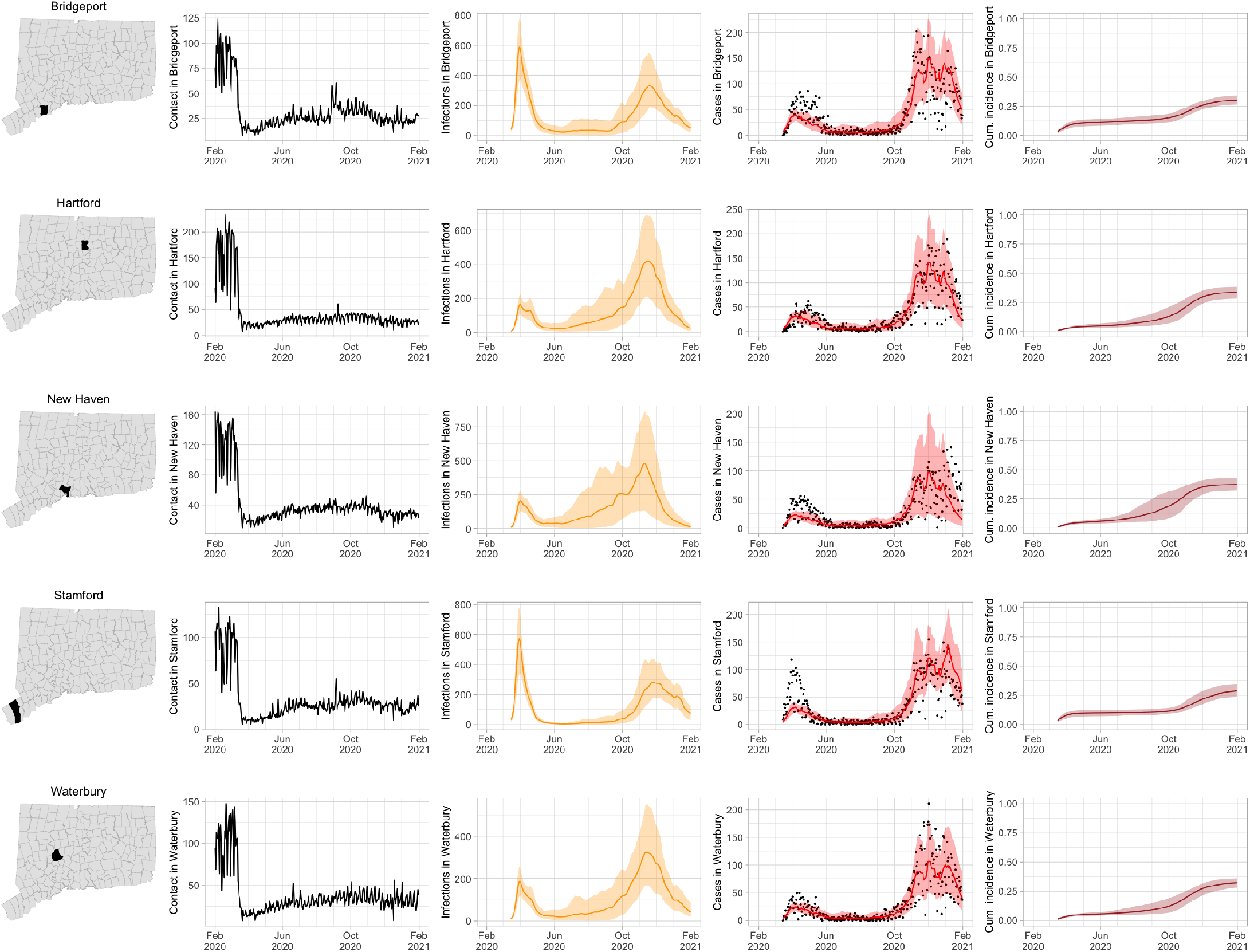
Contact rates, COVID-19 cases, and model predictions (with 95% uncertainty intervals) of infections, cases, and cumulative incidence proportion in the five largest cities by population in Connecticut: Bridgeport, New Haven, Hartford, Stamford, and Waterbury. Black dots show confirmed non-congregate COVID-19 case counts.

### Role of contact in local outbreaks

As COVID-19 case counts in Connecticut decreased during June–August, new and more heterogeneous patterns of transmission emerged. Figure S4 shows contact rates, confirmed non-congregate COVID-19 case counts, and 95% uncertainty intervals for cases in five Connecticut towns where incidence patterns differed from those of the larger cities shown in Figure S3.

**Figure 4:**
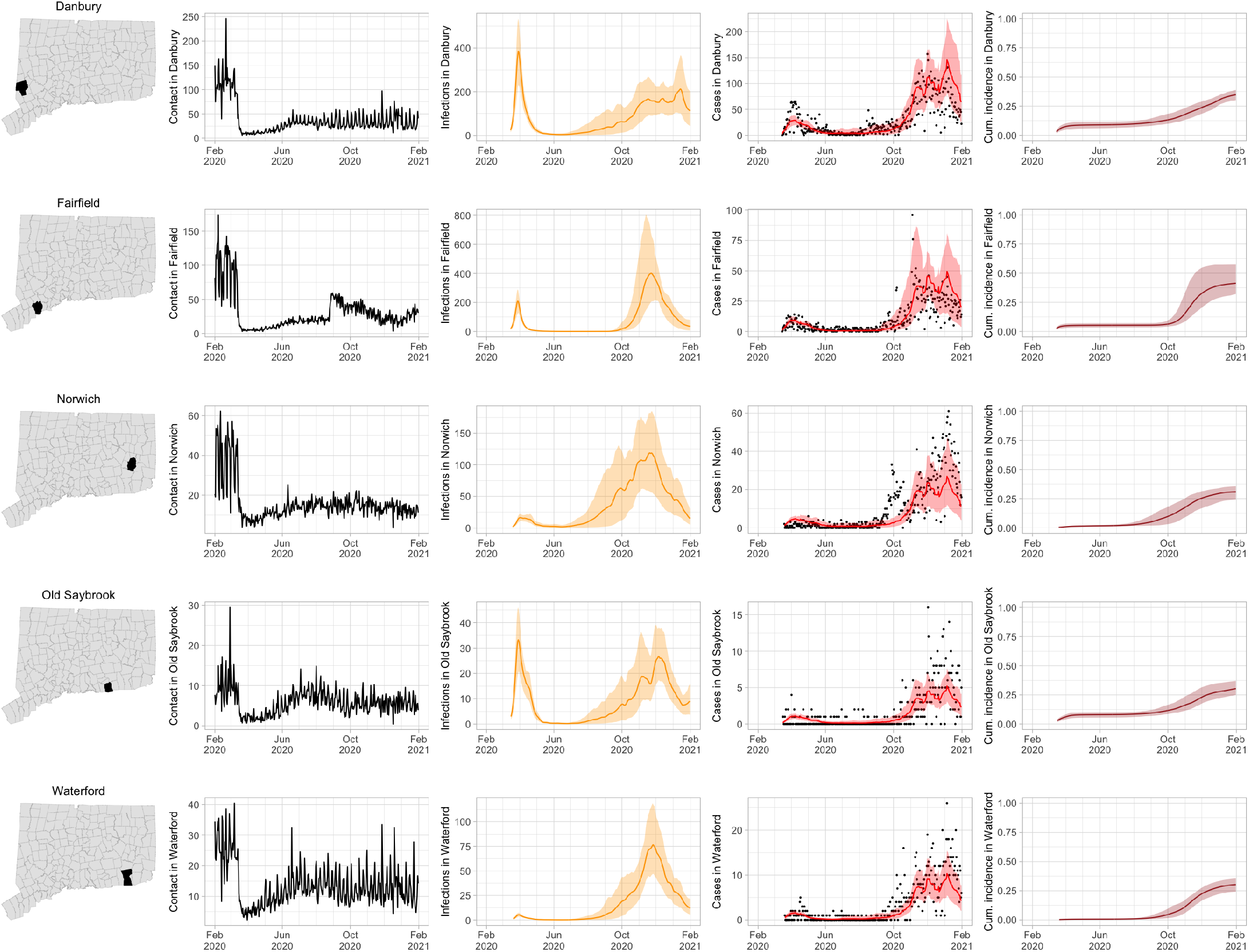
Contact rates, COVID-19 cases, and model predictions (with 95% uncertainty intervals) of infections, cases, and cumulative incidence proportion in several towns in Connecticut whose case or contact patterns differ from that of the state as a whole: Danbury, Fairfield, Norwich, Old Saybrook, and Waterford. Public health officials declared an outbreak in Danbury in mid-August 2020. Fairfield experienced outbreaks linked to two universities in September 2020. Norwich, Old Saybrook, and Waterford, in the eastern part of the state, were mostly spared during the first wave of infection, and had quickly rising case counts in fall 2020.

During June–August, the only known community-wide COVID-19 outbreak in Connecticut occurred in the town of Danbury (population 84,479) [52]. During August 2–20, at least 178 new COVID-19 cases were reported, a significant increase from 40 cases reported during the prior week. Contact tracing investigations by public health officials attributed the outbreak to travel, but the contact rate was high in Danbury beginning in July and genomic analyses suggested the outbreak was closely linked to lineages already circulating in New York City and Connecticut [75, 76]. Predictions from the model including contact rates from Danbury suggest that this outbreak might have been part of a long-term increase in infections that began earlier in July and continued mostly unabated through November.

The town of Fairfield, bordering the larger city of Bridgeport, has a population of 62,105 people, and contains two universities, both of which reopened for in-person education in mid-August. The university communities experienced a surge in cases during September–October after students returned [77]. Students had access to frequent COVID-19 testing, and test coverage in this community was likely higher than in the general population, so infections among students might have been more likely to be reported to public health authorities. Contact rates in both Fairfield and the adjacent city of Bridgeport increased (Figures S3 and S4) during September shortly after students arrived on campus. The consequence of this increase in contact rate is evident in the rise in case counts for Fairfield two to three weeks later.

The eastern part of Connecticut was largely spared in the first wave of infections during March–April, but Norwich (population 39,136) and nearby towns experienced a strong surge in cases beginning in mid-September [54, 55]. Contact rose more quickly in these towns, compared to the western part of the state, following the beginning of Phase 1 in May 2020. Low testing coverage during the spring and summer of 2020, imported infections from neighboring Rhode Island, and lower compliance with social distancing measures might have played a role in outbreaks in the eastern part of the state.

Contact data do not explain all variations in confirmed non-congregate COVID-19 case counts. Though the model fits cases well overall in large cities, it can fail to capture variation in case counts in smaller cities where testing coverage is lower, or in settings where case-finding effort varied over the time. For example, high case counts corresponding to outbreak investigations involving extensive testing in Danbury during August, and Norwich during September/October, do not directly reflect changes in contact, and are not captured by the model projections.

### Contact may provide advance warning of COVID-19 cases

To assess the relationship between close interpersonal contact and COVID-19 cases without SEIR-type model assumptions about the dynamics of transmission, we fit a hierarchical Bayesian space-time statistical model to predict cases using town-level contact data. In a model that included 28 prior days of contact data, lagged contact from 3 to 7 days prior is significantly associated with current-day cases, in agreement with known features of the time to development of symptomatic disease [78–83]. An increase of 10 contacts in each of the previous 28 days within an average town gives rise to an increase in cases by a factor of 1.29 (95% credible interval [1.22, 1.37]) within that town. A model that includes contact predicts cases better than one without contact, according to goodness-of-fit criteria [84, 85]. The model structure and results are described in detail in the Supplementary Materials.

## Discussion

Public health decision-makers track the COVID-19 pandemic using metrics – syndromic surveillance data, cases, hospitalizations, deaths – that lag disease transmission by days or weeks. In this paper, we have described a method for population-level surveillance of close interpersonal contact, the primary route for person-to-person transmission of SARS-CoV-2, by using anonymized mobile device geolocation data. The contact rate can reveal high-contact conditions likely to spawn local outbreaks, or areas where residents experience high contact rates, days or weeks before the resulting cases are detected by public health authorities through testing, traditional case investigation, and contact tracing. Because mobile device geolocation data are passively collected, contact rates are invariant to allocation and availability of public health resources for case finding. For this reason, contact rates could serve as a better early-warning signal for outbreaks than cases alone, especially when test volume is low. Contact rates could also have advantages over surveillance approaches using mobility metrics because interpersonal contact within six feet is more directly related to the likelihood of disease transmission by direct contact or respiratory droplets.

Contact rates could benefit public health efforts to prevent transmission of SARS-CoV-2 in two ways. First, community engagement programs could be directed to locations where the contact rate is high to improve social distancing practices or provide additional protective measures like ensuring adequate ventilation, environmental cleaning, and mask use. Second, enhanced testing in areas with high contact rates, and residential areas of people experiencing that contact, could lead to earlier and more complete detection of cases. Earlier and more complete detection of cases enables faster and more complete isolation of cases and quarantine of contacts, which are crucial to stop transmission and stop outbreaks.

Contact rates also might be a useful addition to mathematical models of infectious disease transmission for prediction of COVID-19 infections or cases. In the early stages of the COVID-19 pandemic, researchers employed variations on the classical SEIR epidemic model [86, 87] to predict the initial wave of infections, estimate parameters like the basic reproduction number, and assess the effect of non-pharmaceutical interventions [e.g. 65, 88, 89, in Connecticut]. These models often assumed a constant population-level contact rate that is subsumed into a transmissibility parameter, or estimated contact rate from survey data collected prior to the pandemic [90, 91].

We have focused in this study on the U.S. state of Connecticut, but the usefulness of anonymized and passively collected contact data could be generalized to other settings. In the U.S., where mobile phone usage is high, states or towns can implement contact surveillance at low cost by working with private sector mobile device data providers. Like Connecticut, other states and countries experienced constrained testing availability in the early stages of the pandemic, and uneven geographic distribution of testing after test volume increased. Non-pharmaceutical interventions such as stay-at-home mandates, business and school closures, and social distancing guidelines also had uneven adoption and compliance varied across time and geography. Surveillance of contact rates could help officials better distribute testing resources and monitor intervention compliance in numerous settings. Internationally, mobile phone ownership has grown quickly but might be low in some developing countries [92], making contact surveillance less feasible in these settings.

The contact rate employed here has several advantages over existing mobility metrics and measures of mobile device density and proximity. First, the contact rate has been designed specifically to measure interpersonal contact within 6-feet relevant to COVID-19 transmission, as defined by CDC [1]. In contrast, mobility metrics primarily measure movement, which might not be a good proxy measure of close interpersonal contact. For each potential contact event between two devices, we use the reported device locations and horizontal uncertainty measurements to compute the probability that the devices were within six feet of one another. In this way, each potential contact event is weighted by the likelihood that the people carrying the devices were close enough for transmission to occur. In contrast, Unacast’s “human encounters” metric measures the frequency of two devices being within 50 meters of one another. Because the Unacast definition includes interactions that are at a distance much farther than six feet, many are unlikely to involve the potential for disease transmission. The contact rate used here incorporates close interpersonal contact occurring in every location in Connecticut, not only at pre-selected venues [e.g. 32, 93] therefore, the contact rate might be a better proxy for population-level transmission risk when there are prevalent infections.

Contact data derived from mobile device geolocation data have limitations. First, not all devices in Connecticut appear in the sample: during May 1– November 28, 2020, we observed a total of 788,842 unique device IDs, representing roughly 22% of the 3.6 million residents of Connecticut. An analysis in the Supplementary Materials shows that there is no evidence of systematic under-coverage of mobile devices as a function of town population sizes, but coverage declines slightly in towns with higher percent of residents identifying themselves as non-White, lacking a high school degree, and below the poverty level. Under-coverage among particularly vulnerable populations could result in under-counting of potential transmission events likely to affect these populations. Second, horizontal uncertainty varies by device and location, making close interpersonal contact that occurs in some areas more difficult to detect with certainty. Third, the duration of time a device was stationary is unknown because location data are reported asynchronously and at irregular intervals. Fourth, using anonymized mobile device geolocation data we do not observe individual-level demographic information, whether a potential contact occurred indoors or outdoors, nor additional individual-level infection risk factors or risk mitigation behavior like mask-wearing, hand washing, avoidance of touching surfaces or avoidance of crowded indoor spaces. However, CDC recommends the determination of close contact should be made “irrespective of whether the person with COVID-19 or the contact was wearing a mask” [94].

The contact rate might not detect all types of close interpersonal contact relevant for disease transmission and does not distinguish between physical contact and close proximity. We exclude contact occurring at primary dwell locations, so contact between pairs of people while at their shared same primary dwell locations is not represented in the contact rate. As a result our model projections may not adequately capture household transmission. Close contact that occurs while traveling, for example on a bus or train, might not be detected because devices are not stationary. Devices located on different floors of the same building might report nearby locations, even if the devices are separated by one or more floors. Location information might be reported by each device at irregular intervals, so we might not observe some kinds of fleeting contact. Contact that occurs outside of Connecticut is not recorded in our dataset. In particular, we did not observe information about contact for people who live in Connecticut and work in the New York City area.

Statewide contact rate based on mobile device geolocation data helps explain Connecticut’s success in avoiding a broad resurgence in COVID-19 cases during June–August 2020, emergence of localized outbreaks during late August–September, and a broad statewide resurgence during October–December. In addition to explaining historical patterns of transmission, incorporating contact rates into an SEIR transmission model might improve prediction of future COVID-19 cases and outbreaks at the town level, which could inform targeted allocation of public health prevention measures, such as SARS-CoV-2 testing and contact tracing with subsequent isolation or quarantine. Contact rate estimated from mobile device geolocation data can help improve population-level surveillance of close interpersonal contact, guide public health messaging campaigns to encourage social distancing, and in allocation of testing resources to detect or prevent emerging local outbreaks.

## Data Availability

All data required to reproduce the analysis are available from the authors.

## Acknowledgements

We are grateful to Gary Archambault, Anderson Brito, Maciej F Boni, Deidre Gifford, Gregg Gonsalves, Nathan Grubaugh, Edward Kaplan, Albert Ko, Jessica Lee, Sean Nolan, Saad Omer, Jasjeet Sekhon, Olivia Schultes, Mary Souther, Lynn Sosa, Robert Tollefson, and Robert Zalot. We thank Stony Brook Research Computing and Cyberinfrastructure, and the Institute for Advanced Computational Science at Stony Brook University. We thank X-Mode Social for providing the source data used to develop the contact metrics. We are grateful fo aggregated mobility data provided by Google, Apple, Facebook, Descartes Labs, and Cuebiq. The findings and conclusions in this report are those of the authors and do not necessarily represent the official position of the CDC.

## Authors contributions

Conceptualization: FWC, JB, SJ; Methodology Development: FWC, OM, ZRL, JLW, JB, JC, PK, TV; Software Development: FWC, OM, ZRL, SJ, JLW, SGD, JC, PK, TV; Validation: FWC, OM, SGD; Formal Analysis: FWC, OM, JLW; Resources: FWC, JB, MC; Data Curation: SJ, OM, JB, JC, PK, TV; Writing – Original Draft: FWC; Writing – Review & Editing: FWC, SJ, OM, ZRL, SGD, JLW, TV, JC, JB, MC; Visualization Preparation: FWC, SGD, OM, ZRL; Supervision Oversight: FWC, JB, MC; Project Administration: FWC, JB; Funding Acquisition: FWC, JB, MC.

## List of Supplementary Materials

Materials and Methods

Supplementary Text

Table S1–S3

Fig S1–S32

References

## Supplement

## 1 Materials & Methods

### 1.1 Overview: computing the contact rate

Anonymized mobile device geolocation data for devices located within the state of Connecticut are received from a third-party provider, X-Mode. Mobile device users consent to sharing of their location information when they accept the terms and conditions of the applications that report location data. Mobile devices also permit users to turn off location sharing. The mobile device geolocation records contain anonymized unique device IDs that persist over time, GPS coordinates, date/time stamps, and GPS location error estimates (also called horizontal uncertainty) measured in meters. The data used in the analysis presented in this paper contain no individually identifying meta-data.

From these records we calculate the location in which each device had the most location records and designate that area as the device’s primary dwell location. The primary dwell location is used to determine whether distinct devices reside at the same location. A device’s primary dwell location is considered to be the best estimate of the home location of the device user. We do not use time of day to determine primary dwell location, as this could lead to mis-characterization of primary dwell location for people who work at night instead of during the daytime, for example.

For each device, we identify clusters of records as locations where other devices were stationary and nearby. When two devices were stationary and in proximity to one another at the same time, the device locations and their corresponding GPS location errors are used to calculate a probability that the devices were in close contact, within six feet (approximately two meters). The resulting information is used to generate a contact event record including the date, time, location and horizontal accuracy of each device involved in the contact, both device IDs, and the computed probability of contact between the devices.

To avoid measuring spurious contact between people who are not actually close to one another, or contact between people who live together, we do not record contacts that occur in some places. A buffered polygon derived from roadway center lines is used to determine if a given contact event occurred on a roadway. If so, then the contact record excluded from the calculation of the contact rate within that region. This could result in missing close contact that occurs in vehicles, such as on buses, trains, or carpooling. Similarly, all contact events for devices at their estimated primary dwell location are tagged and excluded when computing contact rates.

### 1.2 Measuring close contact

We consider a simplified version of geospatial location information on Euclidean plane ℝ^2^. We modify this setting below to account for the curvature of the earth. Suppose that for device location point *i* we observe the triple (*X*_*i*_, *Y*_*i*_, *R*_*i*_) where (*X*_*i*_, *Y*_*i*_) is the reported location (in longitude and latitude) of the mobile device and *R*_*i*_ is the radius of horizontal uncertainty associated with the device location. We assume that the horizontal uncertainty radius *R*_*i*_ is the (1 − *α*) × 100% quantile of the radial density of the device location. Specify this distribution as a symmetric bivariate Gaussian centered at the true device location (*µ*_*x*_, *µ*_*y*_) with covariance matrix 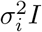, where *I* is the 2 × 2 identity matrix. Then (*X*_*i*_, *Y*_*i*_) has density

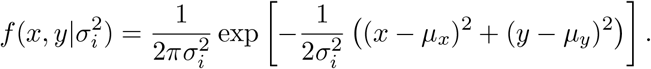

If *R*_*i*_ = *r*_*i*_ is the horizontal uncertainty associated with the (1 − *α*) × 100% quantile radial density level set of the point *i*, then *r*_*i*_ = *σ*_*i*_Φ^−1^(1 − *α*), where Φ^−1^(·) is the standard normal quantile function. We can therefore estimate the variance 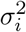 by

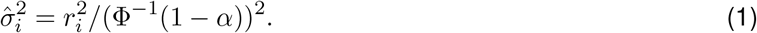

In this paper, we use *α* = 0.05.

Define the Euclidean distance between the reported location of two points (*X*_*i*_, *Y*_*i*_, *R*_*i*_) and (*X*_*j*_, *Y*_*j*_, *R*_*j*_) as

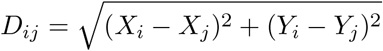

and fix a distance *ϵ* > 0. In this paper, we use *ϵ* equal to two meters. We want to evaluate the probability that points *i* and *j* are within *ϵ* meters of one another. This probability can be expressed as

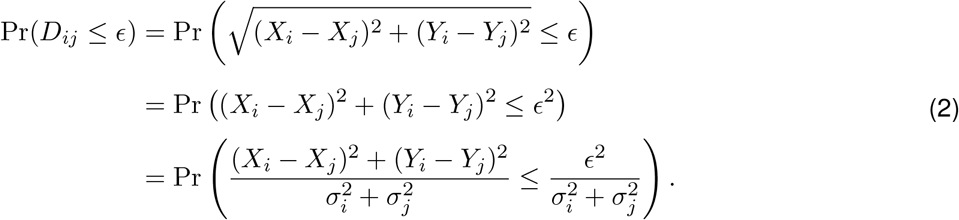

Now under the assumption that (*X*_*i*_, *Y*_*i*_) and (*X*_*j*_, *Y*_*j*_) have independent bivariate Gaussian distribution, the variance-scaled quantity

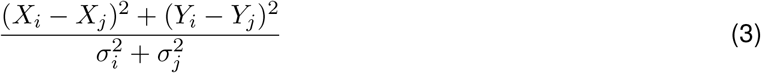

follows the non-central chi-square distribution with 2 degrees of freedom and non-centrality parameter

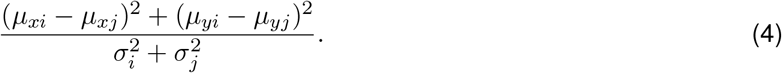

Since the true device locations and variances in (4) are not observed, we substitute the observed device locations *X*_*i*_, *Y*_*i*_, *X*_*i*_, and *Y*_*j*_, as well as the estimated variances 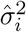 and 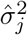 computed from (1). Because the variance-scaled squared distance (3) follows the non-central Chi-square distribution, the probability that the two devices are within two meters, *D*_*ij*_ ≤ 2, can be computed using standard statistical software.

In reality the Earth is not a plane and the Euclidean distance *D*_*ij*_ is shorter than the true distance between *i* and *j* on the surface of the Earth. But for distant points or those whose uncertainty radius is large, it is necessary to evaluate longer distances on the surface of the Earth. We therefore substitute the Haversine distance [1] for the Euclidean distance *D*_*ij*_ in the calculation above. The resulting Gaussian approximation is useful for small geodesic distances because we are interested in points that are less than two meters apart.

To describe computation of the contact rate, let *Z*_*i*_(*t*) = (*X*_*i*_(*t*), *Y*_*i*_(*t*), *R*_*i*_(*t*)) be the location and corresponding horizontal uncertainty radius for device *i* at time *t*. A *potential contact* between devices *i* and *j* at time *t* occurs when the locations of the two devices *Z*_*i*_(*t*) and *Z*_*j*_ (*t*) are stationary and nearby. Let *D*_*ij*_ (*t*) be the computed distance between the two points *i* and *j*. When a potential contact occurs between *i* and *j* at time *t*, let

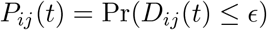

be the probability that these devices are within *E* meters of each other. Let *A*_*ad*_ be the set of pairs of devices for which a potential contact event occurred within area *a* on day *d*. For a potential contact between a pair {*i, j*}, let *t*_*ij*_ be the time of the potential contact. In area *a* on day *d*, the expected number of contacts is the sum of the probabilities of contact, across every potential contact event. We compute two contact rates for each area *a* and day *d*. First, we aggregate contact probabilities by the area in which the contact occurred. The contact rate by region of contact is

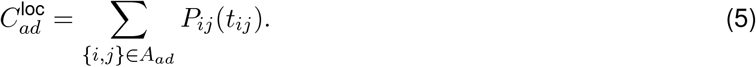

Next, we aggregate contacts by the region (town) of the device’s primary dwell location. Let *A* be the set of all regions and let *h*(*j*) be the primary dwell region of device *j*. The device home contact rate is

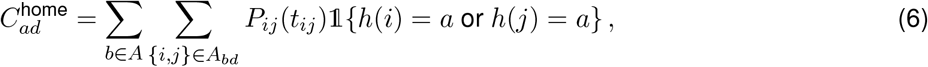

where the indicator function 𝟙{·} is 1 if its argument is true, and 0 otherwise.

### 1.3 Community case definition

Laboratory tests for SARS-CoV-2 and cases of COVID-19 are reportable to the Connecticut Department of Public Health [2]. A confirmed case of COVID-19 is defined as detection of severe acute respiratory syndrome coronavirus 2 ribonucleic acid (SARS-CoV-2 RNA) in a clinical or autopsy specimen using a molecular amplification test [3]. In this analysis, COVID-19 cases and SARS-CoV-2 tests reported to the Connecticut Department of Public Health among persons residing in long-term care facilities, managed residential communities (e.g., assisted living facilities) or correctional institutions were excluded. Cases and tests among staff working at these locations were not excluded.

The Connecticut Department of Public Health used data from COVID-19 case report forms and laboratory test reports to identify cases and tests among residents of congregate settings as follows. For COVID-19 cases, public health follow-up is dependent on whether the person resides in a congregate setting (facility does contact tracing) or not (local and state public health do contact tracing). Therefore, the Connecticut COVID-19 case report form includes a field for healthcare providers to indicate whether the patient resided in a congregate setting, and if so, the type of setting. Because case report forms are not received for all cases, this information might also be completed during case investigations by state or local health department staff. For this analysis, cases among congregate setting residents were excluded by using these case report form data.

To identify tests among congregate setting residents, the Connecticut DPH used a geocoding process because residence in a congregate setting is not included in the laboratory result report form, and the number of tests is too large for public health staff to review individual records. This means that the likelihood of misclassification of congregate setting residence among cases is lower than the likelihood of misclassification for tests.

The home address of each person tested is geocoded by the Connecticut Department of Public Health by using a custom composite locator that first attempts to match on the parcel, then street map data. Approximately 90% of test reports are successfully geocoded. To identify tests performed among residents of congregate settings, geocoded results are compared to a spatial layer of known long term care facilities, managed residential communities, and correctional institutions. Records geocoded to within a 150 foot buffer around these locations are categorized as tests performed among residents of congregate settings.

After the steps above were used to exclude cases and tests among congregate setting residents, the Connecticut Department of Public Health tabulated the aggregate count of cases and tests by town for each day, which were used in this analysis. No individually identifiable information was used in the analysis reported here, nor were any linkages made between mobile device data and cases and tests reported to the Connecticut Department of Public Health.

### 1.4 Data privacy and confidentiality

The mobile device location data contain no individually identifying meta-data. After computing the probability of contact for each potential contact event, we sum contact probabilities by day and by town using (5) and (6). No individual device geolocation data are retained or used in the analysis presented in this paper.

### 1.5 Transmission model for prediction of COVID-19 infections, cases, hospitalizations, and deaths

We developed a county-stratified deterministic model based on the susceptible-exposed-infective-removed (SEIR) framework to represent COVID-19 transmission and predict case counts in Connecticut using the contact rate. Here we provide a brief description of the model and calibration approach. A detailed description of the model is given in [4]. In the model, the population of each county is divided into 10 compartments. Individuals begin in the susceptible (*S*) compartment. Exposed individuals (*E*) may develop either asymptomatic (*A*), mild (*I*_*M*_), or severe (*I*_*S*_) infection. Asymptomatic and mild infections resolve without hospitalization and do not lead to death. Mild symptomatic cases self-isolate (move to *R*_*M*_) shortly after development of symptoms, and transition to recovery (*R*) when infectiousness ceases. All severe cases require hospitalization (*H*) unless hospitalization capacity is exhausted, in which case they transition to 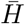 representing hospital overflow, then to recovery (*R*) or death (*D*). The model assumes a closed population and does not capture non-COVID-19 deaths. Let *N*_*i*_ be the population size of county *i* and *J*_*i*_ the set of counties adjacent to county *i*. Let *C*^(*i*)^ represent hospitalization capacity in county *i*, which may vary over time. Transmission dynamics for county *i* are given by the following system of ordinary differential equations (ODE):

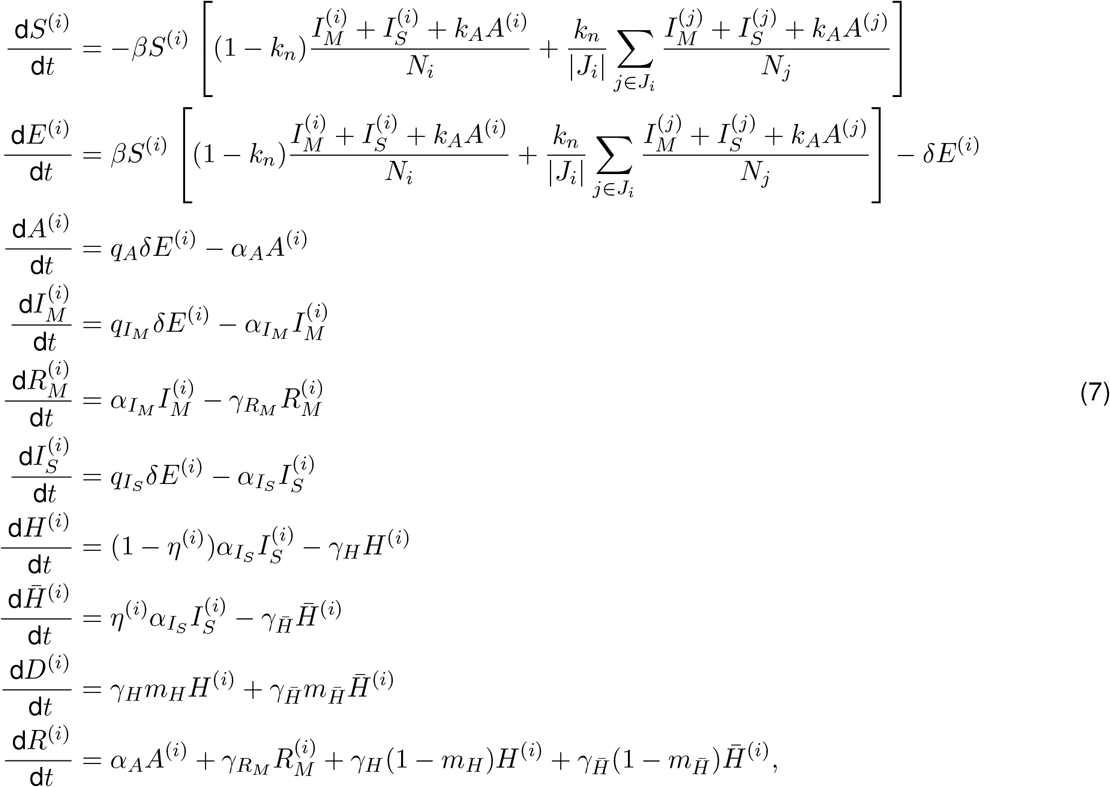

Where 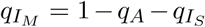. The function 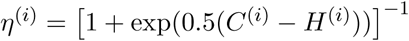 is a “soft” hospitalization capacity overflow function. The models captures infection “community” transmission in non-congregate settings, and excludes cases and deaths occurring in settings like nursing homes and prisons. We assume that recovered individuals remain immune to reinfection for the duration of the study period. The analysis was performed using the R statistical computing environment [5]. We used package deSolve to perform numerical integration of the ODE system [6].

#### 1.5.1 Time-varying model parameters

Recognizing that many of the model parameters are unlikely to be constant over time, we represent the most critical parameters as functions of time. These parameters include: transmission parameter *β*, rates 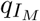 and *α*_*A*_ (reciprocals of average duration of infectiousness among mildly symptomatic and asymptomatic individuals), 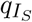 - proportion of infections, which are severe and require hospitalization, rate of hospital discharge *γ*_*H*_, and hospital case fatality ratio *m*_*H*_.

We use data on close contact to parameterize temporal dynamics of transmission parameter *β*:

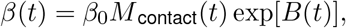

where *M*_contact_(*t*) is a measure of close contact at time *t* relative to the pre-epidemic level, and exp[*B*(*t*)] is a function that approximates residual changes in transmission parameter that are not explained by changes in close contact or other time-varying parameters. *B*(*t*) is a smooth function obtained by applying spline smoothing on a piecewise linear function *B*^*\**^(*t*), where *B*^*\**^(*t*) is modeled with 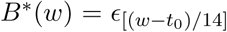 defined on bi-weekly knots *w* = {*t*_0_, *t*_0_ + 14, *t*_0_ + 28, …} over the observation period and linearly imputed between the knots. We model the vector of random effects ***E*** using a random walk of order one:

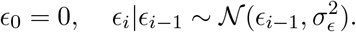

For the hyperparameter *σ*_*ϵ*_ we let

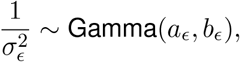

with shape parameter *a*_*ϵ*_ = 2.5 and rate parameter *b*_*ϵ*_ = 0.1.

Rates of isolation or recovery *α*_*I*_*M* and *α*_*A*_ are assumed to depend on the testing volume. Widespread testing is assumed to identify infectious cases sooner, resulting in faster isolation and shorter duration of infectiousness. Via its relationship to age distribution of cases, probability of severe infection is assumed to change over time, likely due to higher adoption of social distancing measures by individuals with worse prognosis [7]. Time-varying rate of hospital discharge *γ*_*H*_ was obtained from the Connecticut Hospital Association (CHA) based on the analysis of insurance claims data. Temporal variation in the hospital case fatality ratio *m*_*H*_ was estimated based on data on daily hospital deaths and admissions.

Table 1 summarizes the parameters in the model and their meaning. We fix some of the model parameters whose values can be estimated based on prior research.

**Table 1:** Transmission model parameters. Fixed parameters are drawn from the cited sources. Calibrated parameters are estimated using the Bayesian procedure described below. Durations are measured in days.

| Parameter | Meaning | Value | Source |
| --- | --- | --- | --- |
| $q_A$ | Proportion of infections that are asymptomatic | Calibrated, prior mean = 0.4 | [8–10] |
| $q_{IS,0}/(1 - q_A)$ | Proportion of infections that are severe among symptomatic | 0.065 | [11–13] |
| $\beta_0$ | Transmission parameter at baseline | Calibrated, prior mean = 1 | Diffuse prior assumed |
| $\delta$ | 1/duration of latency | 1/4 | Of 6 days incubation, 2 days are presymptomatic [10, 12, 14–23] |
| $\alpha_{A,0}$ | 1/duration of asymptomatic infectiousness at baseline | 1/7 | [16, 24–29] |
| $k_A$ | Relative infectiousness of asymptomatic compared to symptomatic | 0.5 | [9, 15, 26, 30, 31] |
| $\alpha_{IM,0}$ | 1/duration of symptomatic infectiousness at baseline | 1/4 | [11, 12, 14]; of 4 days, 2 days are presymptomatic [15–17] |
| $\gamma_{RM}$ | 1/duration of isolation among mild symptomatic | 1/7 | [24, 28, 29, 32] |
| $\alpha_{IS}$ | 1/time between infectiousness onset and hospitalization | Calibrated, prior mean = 1/9 | [15–17, 33, 34] |
| $\gamma_H$ | 1/length of hospital stay | Time-varying | CHA data on average hospital length of stay by month [35] |
| $m_{H,0}$ | Hospital case fatality ratio at baseline | Calibrated, prior mean = 0.13 | Estimated from CHA COVID-19 hospitalization data [35] |
| $k_n$ | Proportion of contact between adjacent counties | 0.015 | Assumed |
| $E_0$ | Number of exposed individuals at the epidemic onset | Calibrated, prior mean = 150 | Assumed |
| $L_H$ | Hospitalization reporting lag | Calibrated, uniform prior on integers between 5 - 14 days | Assumed |
| $L_D$ | Death reporting lag | Calibrated, uniform prior on integers between 5 - 14 days | Assumed |

#### 1.5.2 Calibration data

We calibrate the joint distribution of model parameters at the state level using the observed dynamics of confirmed COVID-19 hospitalizations census, cumulative COVID-19 hospitalizations, and cumulative number of deaths among hospitalized cases, which was obtained from the Connecticut Hospital Association [35]. In addition to close contact data, we use the following data to inform time-varying model parameters: average hospital length of stay by month (source: CHA [35]), testing volume data, and data on the age distribution of confirmed cases (source: Connecticut Department of Public Health (CT DPH) daily reports [36]). We calculated non-institutionalized county-level population and age structure in Connecticut using the 2014–2018 estimates of the American Community Survey [37]. Daily total available hospital beds (including occupied) in each county were obtained from the Connecticut Hospital Association/CHIMEData [35, 38] and used as hospitalization capacity values on a given date.

Since available hospitalizations data does not disaggregate by the patient’s place of residence at the time of diagnosis or hospitalization, we estimate the time-varying distribution of hospitalizations and hospital deaths coming from congregate and non-congregate settings based on the daily distribution of COVID-19 death counts occurring in hospitals by the type of residence at the time of diagnosis (congregate vs. non-congregate) and a case fatality ratio among hospitalized residents of congregate settings. These data were obtained from Connecticut Department of Public Health.

#### 1.5.3 Model calibration and Bayesian posterior inference

We calibrate a joint posterior distribution of model parameters 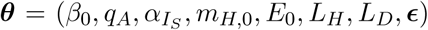 and hyperparameters ***σ*** = (*σ*_*h*_, *σ*_*u*_, *σ*_*d*_, *σ*_*ϵ*_) to the observed data using a Bayesian approach with a Gaussian likelihood. We also accommodate reporting lags in observed hospitalizations (*L*_*H*_) and hospital deaths (*L*_*D*_). The distributions of hospitalizations census, cumulative hospitalizations, and hospital deaths are given by:

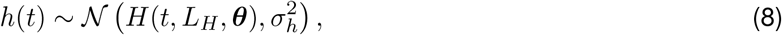

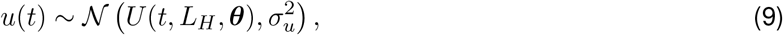

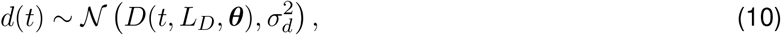

where *H*(*t, L*_*H*_, ***θ***), *U* (*t, L*_*H*_, ***θ***), and *D*(*t, L*_*D*_, ***θ***) are model-projected hospitalizations census (lagged by *L*_*H*_), cumulative hospitalizations (lagged by *L*_*H*_), and cumulative hospital deaths (lagged by *L*_*D*_) at time *t* under parameter values ***θ***. Reporting lags *L*_*H*_ and *L*_*D*_ are correlated with other model parameters, including latency period, time between infection and hospitalization, time between infection and death, and length of hospital stay, therefore *L*_*H*_ and *L*_*D*_ should not be strictly interpreted as reporting lags. We assume the date of epidemic onset to be February 16, 2020 - 21 days before the first case was officially confirmed in Connecticut on March 8th, 2020, and initialize the model with *E*_0_ exposed individuals at the time of epidemic onset and set the initial size of all downstream compartments to be zero. We assume a prior distribution on *E*_0_ and calibrate it along with other model parameters. The county-level distribution of *E*_0_ is fixed and was estimated based on the county population size and dates of first registered case and death in each county. We put the same independent Inv-Gamma(*a, b*) prior on the three hyperparameters 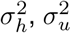, and 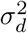 with *a* = 50 and *b* = 5 × 10^7^.

We construct the joint posterior distribution over unknown parameters (***θ, σ***) as

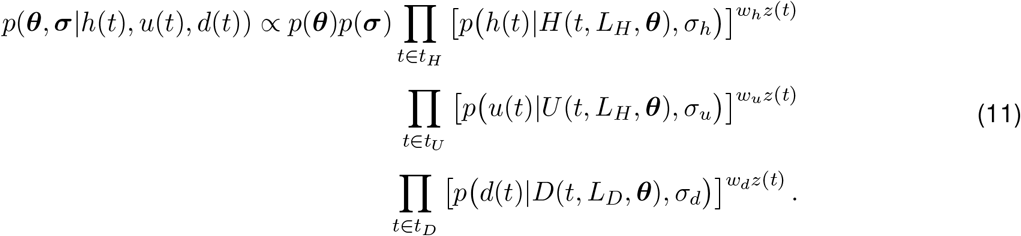

Each likelihood term is weighted by *z*(*t*) (observation weight at time *t*) times the weight assigned to a respective time series. We set *w*_*h*_ = 0.89, *w*_*u*_ = 0.01, and *w*_*d*_ = 0.1. We place a large weight on the hospitalizations census, since this time series is most sensitive to changes in epidemic dynamics, and a small weight on cumulative hospitalizations, since it measures a feature that is related to hospitalizations census. The range of observation times differ for different time series. For hospitalizations census and deaths, observation times start with the first non-zero observation. For cumulative hospitalizations, observation times start on May 29, 2020 when this indicator started being reported routinely. The last observation in all three time series is from February 15, 2021.

Sampling from the joint posterior distribution of (***θ, σ***) given in (11) is performed using Markov Chain Monte Carlo (MCMC). We employ a hybrid algorithm that combines elliptical slice sampling (ESS) [39], Gibbs sampling, and Metropolis-Hastings sampling with random walk proposals. Details of the algorithm implementation are provided in [4]. Bottom row of Figure 1 shows statewide model fit to hospital census, hospital deaths, and total deaths.

**Figure 1:**
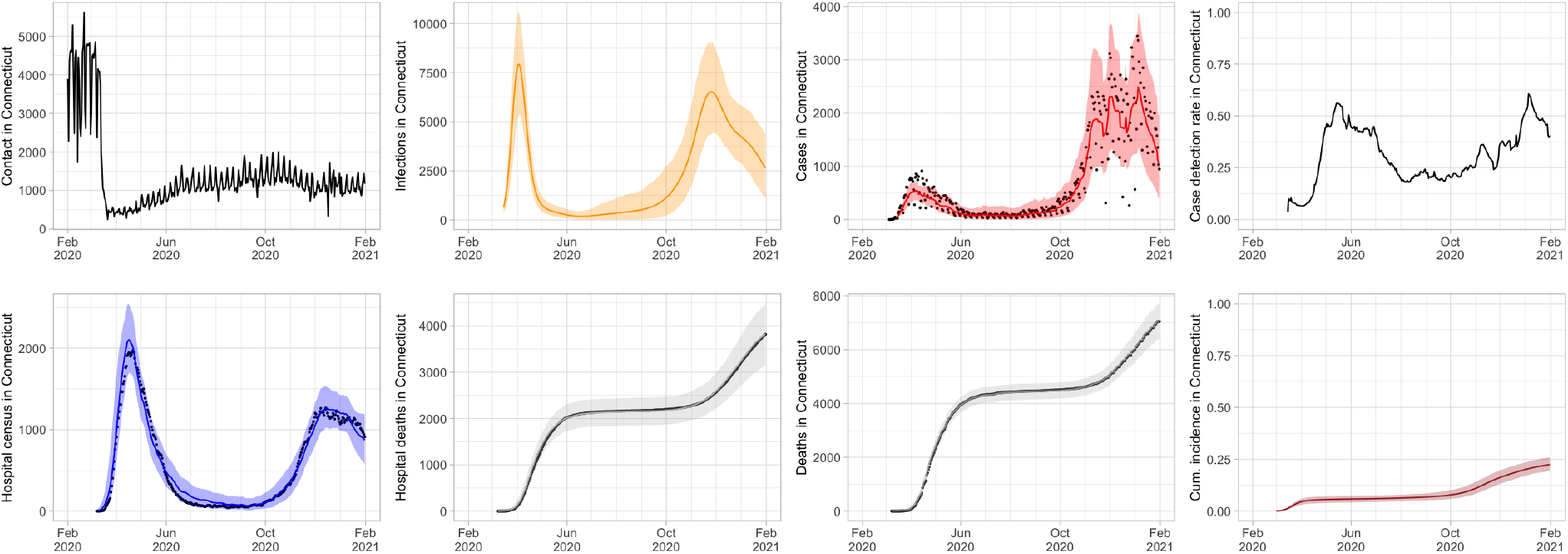
Connecticut statewide data and calibrated model projections. Top row from left to right: statewide contact, projected daily infections, projected (red line) and observed (black dots) cases, estimated statewide case detection rate. Bottom row from left to right: projected and observed hospital census, projected and observed hospital deaths, projected and observed total deaths, estimated cumulative incidence. Model projections are shown as lines with 95% posterior intervals. Observed data is shown as black dots.

#### 1.5.4 Model projections for case counts at the town level

To produce town-level projections, we use parameters estimated from the state-level joint posterior distribution and town-level inputs. Town-level data consist of the normalized close contact function, town population size, and town-specific initial model compartment conditions. Normalized close contact at the town level is obtained by adding an intercept to the town level contact that equals the median difference between the state and the town-level contact for all dates *t* after April 7, 2020. This step enables the application of a random effects vector ***E*** estimated at the state-level to the town-level contact data. The normalization approach preserves town-level contact dynamics and contact levels in each town early in the epidemic and immediately after the implementation of social distancing measures. Town-specific initial conditions were computed from the state-level posterior distribution of *E*_0_ based on the town population size and its proximity to New York City.

We simulate epidemic dynamics for each of the 169 Connecticut towns using the model given in (7), modified to include only data from a single town. Using the model-projected dynamics of lagged daily infections within the town, we estimate the time-varying COVID-19 case-finding rate for each town using a log-linear regression model:

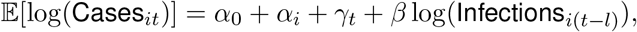

where Cases_*it*_ is the 7-day moving average of reported COVID-19 cases in town *i* at time *t, α*_*i*_ is a town effect, *γ*_*t*_ is a time effect measured daily and Infections_*i*(*t*−*l*)_ is the model-projected number of daily infections in town *i* at time *t* − *l*, and we set *l* = 14 days. As a final step to produce model-based estimates of daily cases in each of 169 Connecticut towns, we truncate regression-estimated town-level case-finding rate at 1 and apply this estimated rate to model-projected daily infections. The top row of Figure 1 shows statewide contact measure, projected infections, estimated and observed non-congregate COVID-19 case counts, and estimated case-detection rate. Estimates of statewide case counts is obtained by summing up estimated cases across 169 Connecticut towns.

### 1.6 Space-time regression model

To assess the relationship between close interpersonal contact and COVID-19 cases without assumptions about the dynamics of transmission, we develop a statistical model for describing the associations between town-level contact in previous days and current day COVID-19 cases using a distributed lag negative binomial regression framework. We emphasize that this “phenomenological” regression model does not use any SEIR-type assumptions about the relationship of contact, infections, and cases. Our purpose is to validate the usefulness of contact data in prediction of future case counts throughout the state.

In the model, each town has a unique set of parameters that describe the lagged associations with past contact, as we hypothesize that there may be spatial differences in these associations due to town-level features (e.g., population density). Spatial correlation between these parameters is also modeled since towns close to each other may exhibit similar spatial patterns. Specifically, the model is given as

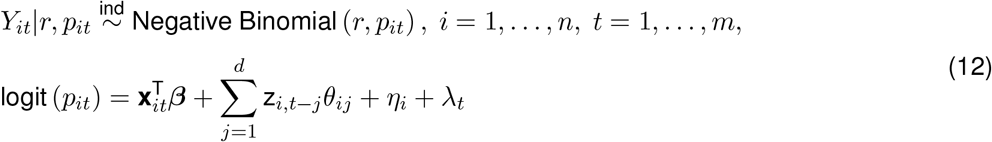

where *Y*_*it*_ is the number of COVID-19 cases recorded in town *i* on day *t, n* is the total number of towns in the analysis (*n* = 169), and *m* is the total number of included days of data (*m* = 351). In our application of the model, the first day of analysis (*t* = 1) occurs on February 29^th^, 2020 and the final day (*t* = 351) occurs on February 13^th^, 2021.

In (12), we use the logit link function to connect the underlying probability that controls the expected number of daily cases in a location to the covariates of interest, where **x**_*it*_ is a vector of town-specific non-time varying covariates (i.e., median household income, total population, percent of the population living below the poverty level, percent of the population that is non-Hispanic White, percent of the population with some college education, and percent of the population that is ≥ 65 years) along with a day-of-the-week indicator to account for systematic case reporting patterns across time; ***β*** is the corresponding vector of regression parameters. Contact in town *i* on day *t* is denoted as z_*it*_, and in our application of the model we include contact up to *d* = 28 days (i.e., 4 weeks) in the past. Town-specific (*η*_*i*_) and time-specific (*λ*_*t*_) random effects are included to account for separable spatial and temporal correlation, respectively, in the case data.

The *θ*_*ij*_ parameters are town- and lag-specific, and describe the association between contact *j* days earlier in the town and current day cases. The collection of all lag parameters from town *i*, ***θ***_*i*_ = (*θ*_*i*1_, …, *θ*_*i*28_)^T^, are decomposed into “global” and “local” effects to account for similarities in associations shared across all towns and town-specific deviations from that trend such that

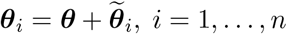

where ***θ*** = (*θ*_1_, …, *θ*_28_) T represents the vector of parameters shared across all towns and 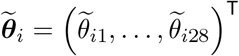, are town-specific deviations from the global pattern. We anticipate that the global distributed lag parameters closer together in lag time will have similar behavior and model this directly by specifying

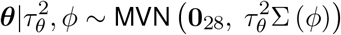

where MVN represents the multivariate normal distribution, **0**_28_ is a vector of 28 zeros, and the correlation between the global lag parameters is defined by

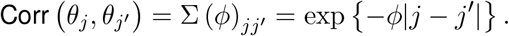

The variance and smoothness of the process are described by 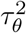 and *ϕ*, respectively, where smaller values of *ϕ* suggest higher correlation between parameters across the lags.

Spatial and cross-correlations between the town-specific lag parameters 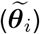 and town-specific intercepts (*η*_*i*_) are modeled using a multivariate conditional autoregressive model [40] given as

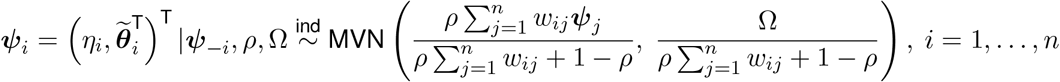

where ***ψ***_−*i*_ is the entire set of ***ψ***_*j*_ vectors with ***ψ***_*i*_ removed and *w*_*ij*_ is an indicator describing whether towns *i* and *j* share a common border or not. As is standard practice, towns are not considered to be neighbors of themselves, resulting in *w*_*ii*_ = 0 for all *i*. This model specifies that *a priori*, the set of parameters from a specific town have a multivariate normal distribution with a mean vector equal to a weighted mean of its neighbors vectors. The amount of spatial similarity between towns is controlled by *ρ* ∈ (0, 1) where a value of *ρ* close to zero indicates near independence and close to one results in strong spatial smoothing. The cross-correlation between parameters within a given town is described by the unstructured covariance matrix Ω.

Finally, *λ*_*t*_ represents a flexible time trend that is common to all towns in the analysis and is defined using an autoregressive model such that

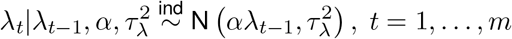

where *λ*_0_ *≡* 0 and *α* ∈ (0, 1) controls the level of similarity across time. This effect serves to account for temporal correlation between the case counts and describe global patterns of risk that vary across time.

#### 1.6.1 Prior distributions

We complete the model by specifying prior distributions for all introduced parameters. When possible, we opt for weakly informative priors so that the data are the main drivers of the inference. The priors are given as *r ∼* Discrete Uniform [1, 20], 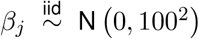 for all 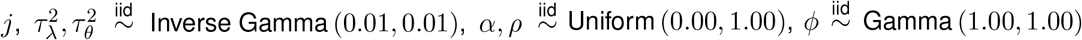, and Ω^−1^ *∼* Wishart (30, *I*_29_) where *I*_29_ is the 29 by 29 identity matrix.

#### 1.6.2 Competing methods

To determine the importance of previous days contact on explaining trends in current day COVID-19 case counts, we also test two competing modeling options. First, we consider the “Global Contact” model, where (12) is modified such that ***θ***_*i*_ is replaced with ***θ*** (***θ*** previously described). As a result, in Global Contact there are no town-specific lag parameters, only a single set that is shared across all towns. We expect this model to perform poorly compared to the full spatio-temporal model in (12) if there is spatial variability between these associations. Next, we consider the “No Contact” model, which modifies (12) by removing all of the contact information entirely. This model is likely to struggle to explain variability in the case data if prior days contact are important predictors. We note that both of these competing models technically represent subsets of the full spatio-temporal model in (12). For example, when ***θ***_*i*_ *≡* **0**, the full model becomes Global Contact and when 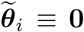 it becomes No Contact. Due to this flexibility, we anticipate that the full model will perform well regardless of the spatio-temporal trends in the data.

We fit each method to the data and compare the findings using multiple methods. First, we use Watanabe–Akaike information criteria (WAIC) [41] to compare the explanatory ability of each method. WAIC is a Bayesian model selection tool that identifies the model, among a small subset of competitors, that has the best balance of fit to the data and complexity, with smaller WAIC values being preferred. Next, we consider a posterior predictive model comparison tool focused on the predictive ability of each method [42]. Specifically, we calculate the posterior mean of the deviance of the negative binomial regression model evaluated at the observed data and data generated from the posterior predictive distribution (from the same design points as the observed data). Smaller values of this metric suggest improved predictive performance of the corresponding method.

#### 1.6.3 Model fitting

We fit all models using Markov chain Monte Carlo sampling techniques, including a Pólya-Gamma latent variable approach for obtaining semi-conjugacy for many of the parameters within the negative binomial regression model [43]. From each model, we collect 100,000 samples from the joint posterior distribution after removing the first 10,000 iterations prior to convergence of the model. We further thin the samples by a factor of 10 to reduce posterior autocorrelation, resulting in 10,000 samples with which to make posterior inference. Convergence of the models was assessed by visually inspecting individual parameter trace plots and calculating Geweke’s diagnostic [44] for all relevant parameters. Neither tool suggested any obvious signs of non-convergence.

## 2 Supplementary Text

### 2.1 Mobile device coverage and representativeness in Connecticut

We investigated the coverage of mobile devices in the sample by town population, percent who do not identify as “only white”, percent without a high school degree, and percent below the poverty level as defined by the federal government’s official poverty threshold, which accounts for household size, number of children, and age of householder. Demographic data were obtained from the American Community Survey [37, 45]. Figure 2 shows that device coverage tracks population size throughout the state, but coverage decreases slightly with the percent non-white, the percent that lack a high school degree, and percent below the poverty level. The town of Union is the smallest town in Connecticut, with a total population of 873 people. We do not know whether lower coverage is due to a lower percentage of town residents owning a mobile device, or because of systematic under-coverage of devices from these towns in our sample.

**Figure 2:**
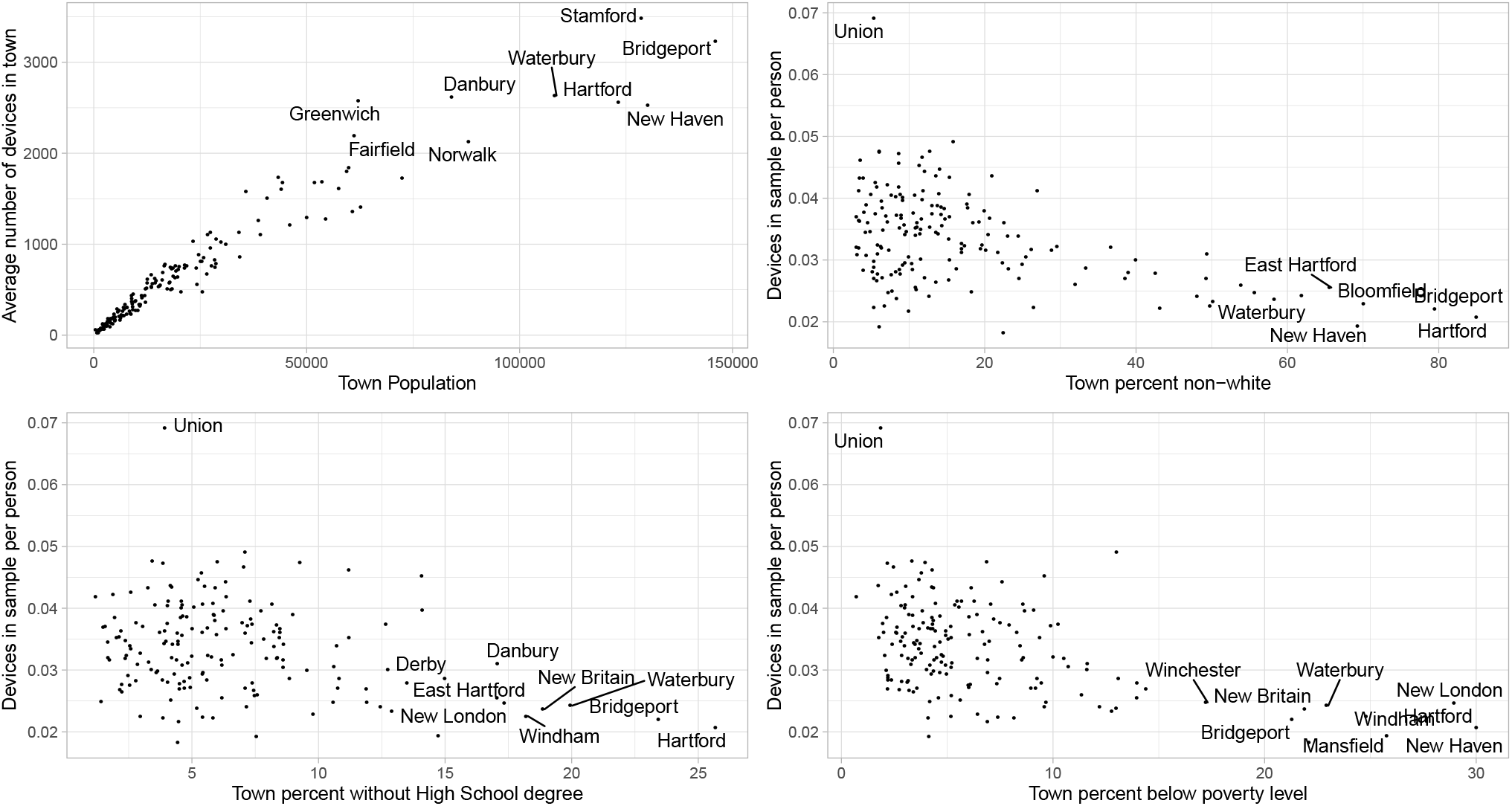
Number of mobile devices in the sample by Connecticut town. Top left: average number of devices in each town by total town population; top right: devices in the sample per person by town percent not identifying as “only white”; bottom left: devices in the sample per person by percent without a high school degree; and bottom right: devices in the sample per person by percent below the poverty level.

We received mobile device geolocation data in two overlapping batches, using different methods of anonymizing device IDs. In the first batch, covering February 1 – May 31, 2020, we observed a total of 573,452 unique device IDs in Connecticut. The average number of unique device IDs per day was 141,617. Each week an average of 80.5% of the device IDs carried over to the next week. In the second batch, covering May 1, 2020 to January 31, 2021, we observed a total of 788,842 unique device IDs in Connecticut. The average number of unique device IDs per day was 114,946. Each week an average of 77.6% of the device IDs carried over to the next week. Connecticut has a population of 3.565 million people [37, 45]. Contacts occurring within a given town are not captured if we cannot calculate a primary device dwell location for both devices involved in the contact. Individual device IDs might drop out of the sample for several reasons:

- the device leaves the state of Connecticut,
- the user uninstalls the applications reporting data to X-Mode,
- the user stops using an application that reports location data only while in use,
- the user rotates their device ID, or
- the user gets a new device with a new advertising ID.

Figure 3 shows the number of unique devices per day in the Connecticut sample.

**Figure 3:**
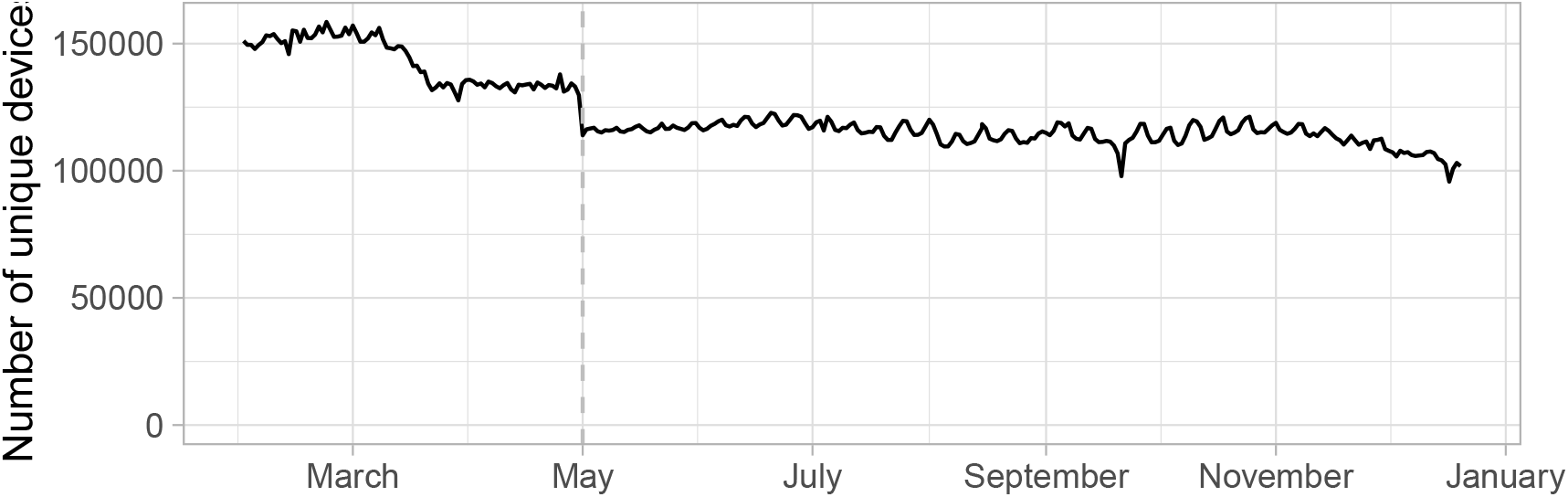
Number of unique mobile device IDs per day in the Connecticut sample. The vertical dashed line on May 1, 2020 corresponds to a change in the unique device ID coding from the data supplier.

### 2.2 Interactive web application

We created an interactive web application to allow users to explore contact patterns in Connecticut over time, available at https://forrestcrawford.shinyapps.io/ct_social_distancing/. Figure 4 shows a screen shot of the interactive application. The application uses the Shiny framework [46] for the R computing environment [5], Leaflet for mapping [47], and tigris for shapefiles [48]. The map explorer is based in part on Edward Parker’s COVID-19 tracker [49] and code [50].

**Figure 4:**
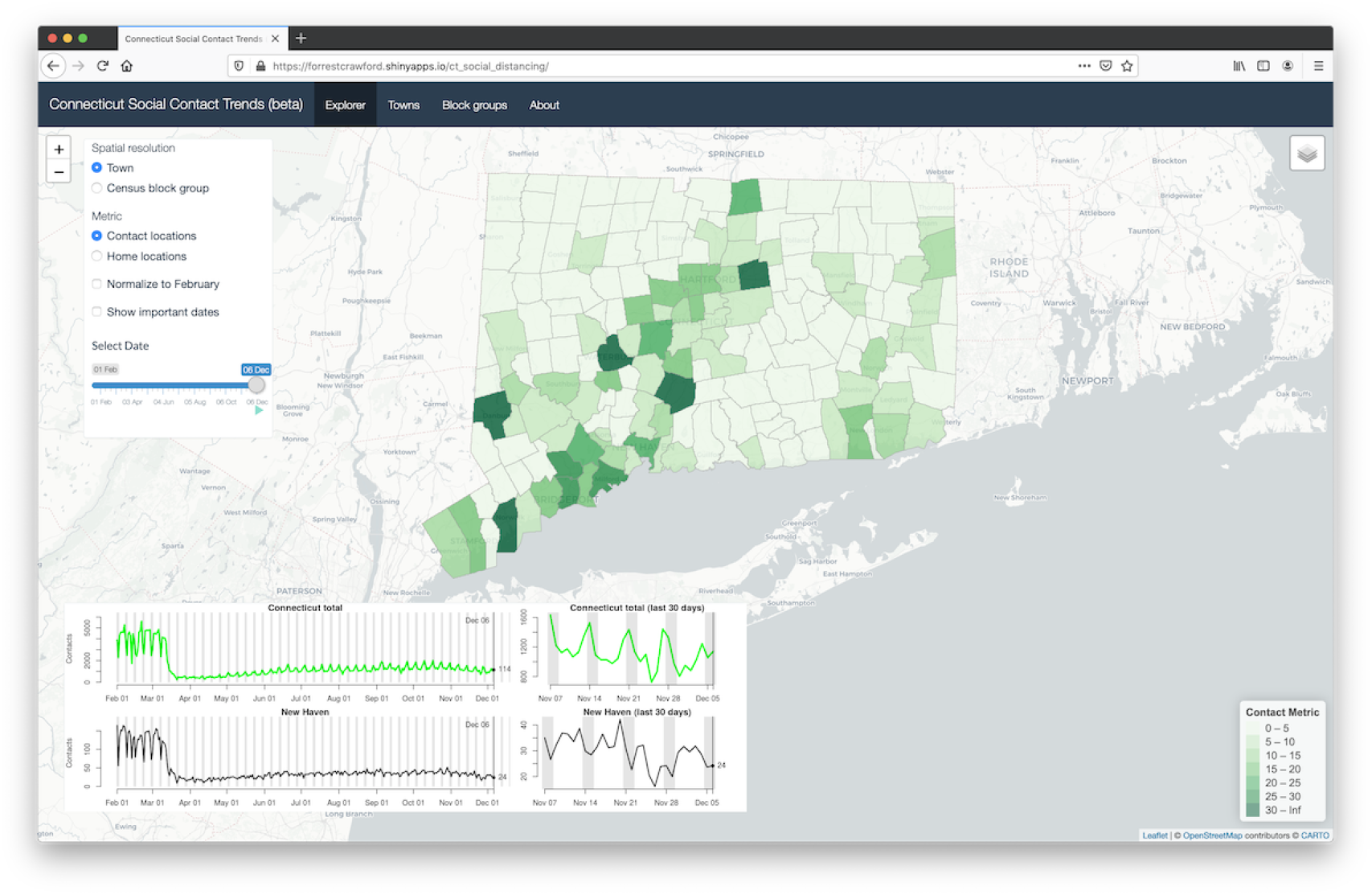
Interactive web application showing contact in Connecticut towns on December 6, 2020. The application can display the locations where contact is occurring or contact by the town of device primary dwell town.

The application shows contact by location of contact (5) and by device primary dwell town 6 for each day, at the town and census block group levels. Users can view the contact maps over time from February 1, 2020 to the present, as well as time trends of contact at the state and local levels. The application shows the top contact towns and census block groups throughout Connecticut, as well as points of interest – businesses, schools, hospitals – that help identify block groups.

### 2.3 Comparison of contact rate to mobility metrics

In order to compare the contact rate described in this paper to other mobility metrics, we acquired Connecticut mobility data from Google [51], Apple [52], Facebook [53], Descartes Labs [54, 55], and Cuebiq [56]. Google and Apple provide public data access, while Facebook and Cuebiq provide data through their respective Data For Good programs. We normalized all metrics to a day-of-week baseline using data from January or February depending on availability, and plot their percent change from baseline from February 2020 through January 2021.

Apple state-level data measures Apple Maps routing requests, categorized as transit, walking, or driving. Map routing requests are a proxy for mobility but might not represent actual trips. Movements for which Apple Maps directions are not needed, such as everyday trips for work, school, or shopping, might not be represented in routing request metrics. Figure 5 shows mobility metrics published by Apple using the day-of-week median during February 2 – February 29, 2020 as a baseline. While transit use remained below baseline during March 2020 through January 2021, driving and walking returned to baseline in June 2020. Driving and walking remained above baseline until November 2020, at which point they returned to near baseline through January 2021.

**Figure 5:**
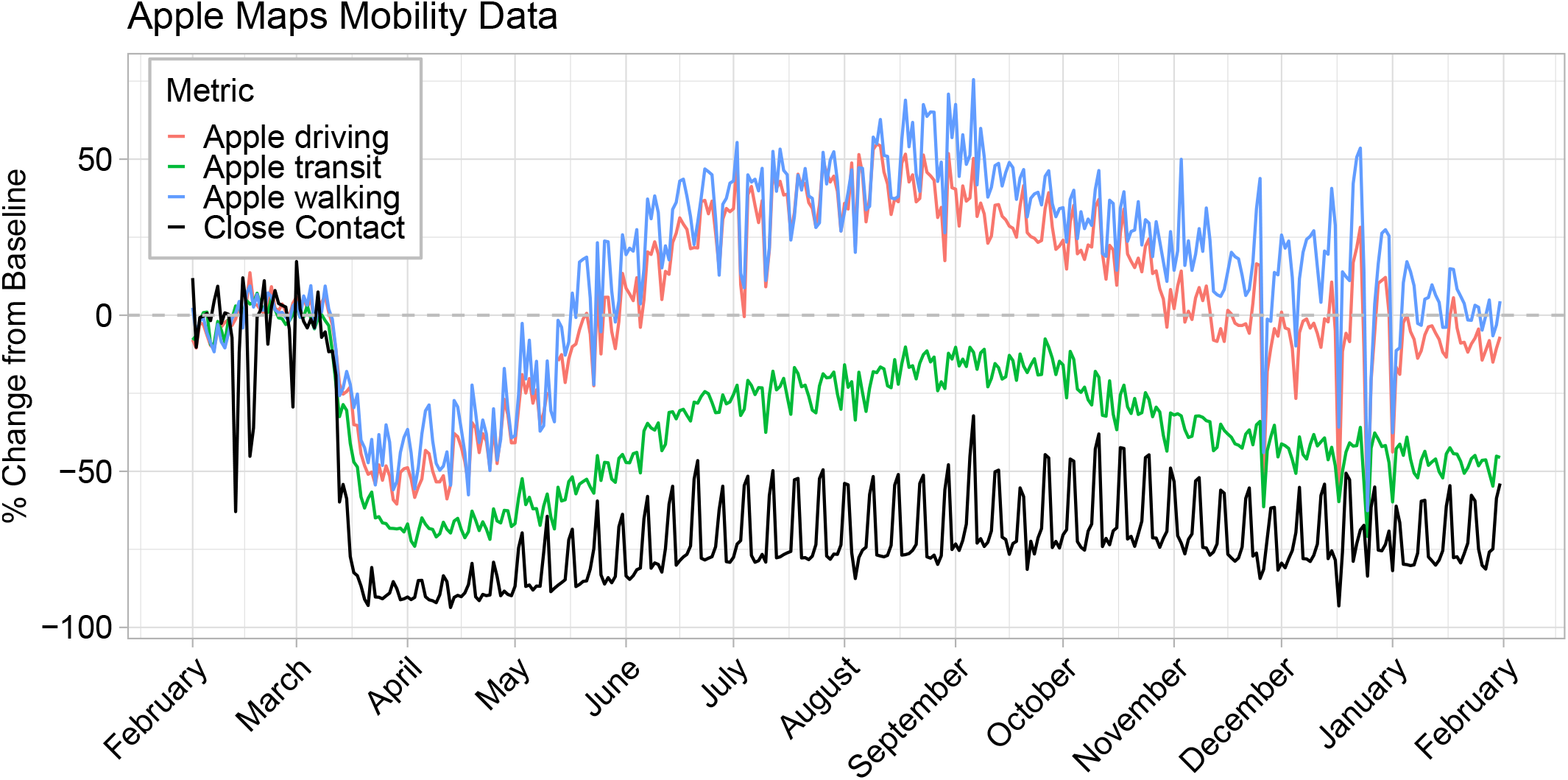
Comparison of Apple Maps mobility metrics to the contactr rate described in this paper during February 1 – January 31, 2021.

Google state-level mobility data measured visits to areas of interest, categorized as grocery and pharmacy, parks, residential, retail and recreation, transit stations, and workplaces. More detailed information about the definitions of these areas of interest, and the completeness of these categories, is not available. Figure 6 shows mobility metrics published by Google using the day-of-week median from January 3, 2020 to February 6, 2020 as the baseline. All categories other than transit stations and workplaces returned to near baseline levels by summer 2020, and all categories other than residential remained near or below baseline throughout winter 2020.

**Figure 6:**
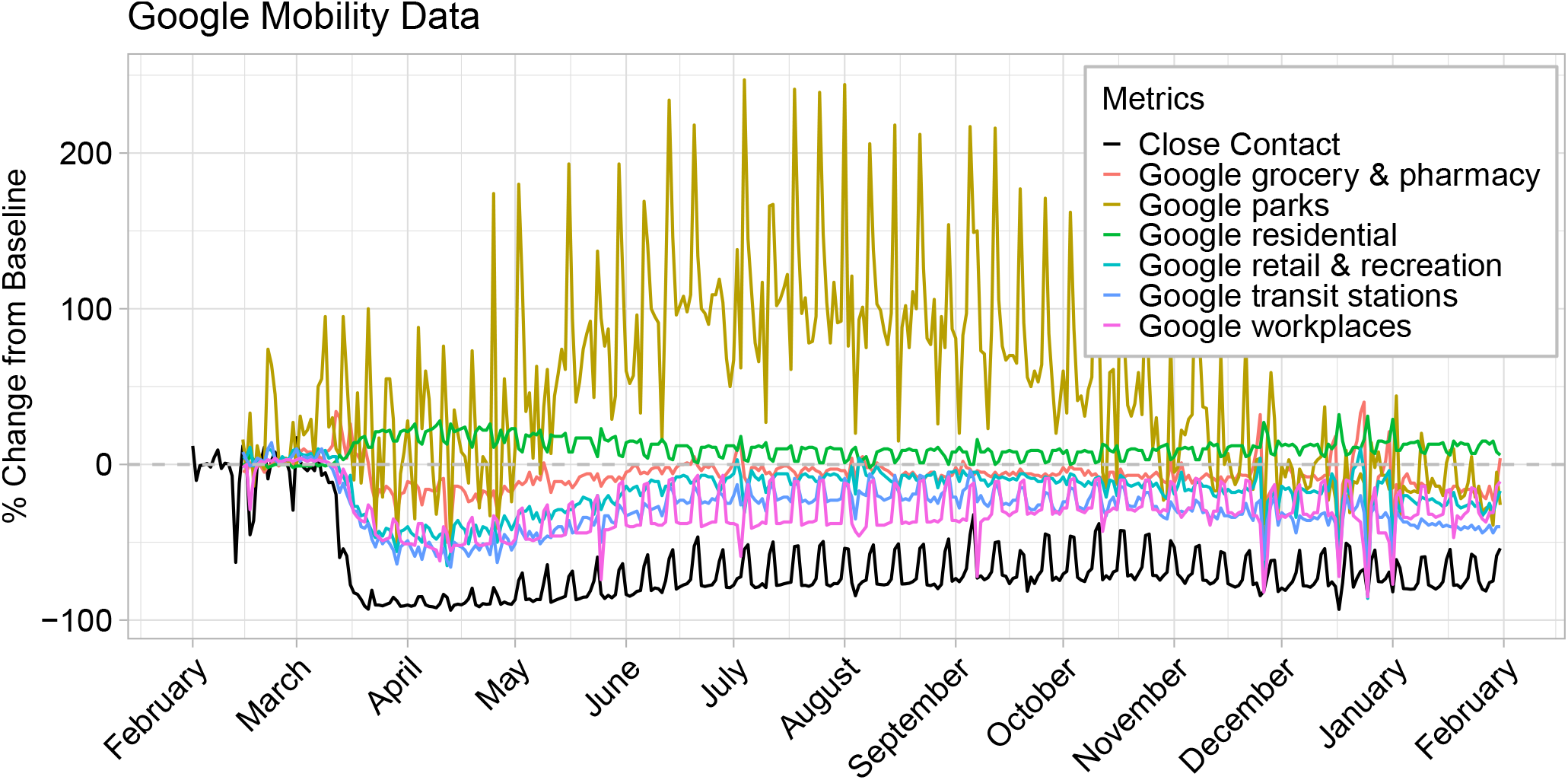
Comparison of Google mobility metrics to the contact rate described in this paper during February 1 – January 31, 2021.

Facebook county-level mobility data measured the number of 600m by 600m geographic units visited by a device in a day. This metric summarizes how mobile people from different counties are, but might not represent the distance of travel, time away from home, or potential close contacts with others. Figure 7 shows mobility metrics published by Facebook with day-of-week mean during February 2 – February 29, 2020 (excluding February 17) as the baseline. Facebook mobility levels returned to near baseline in all Connecticut counties by July 2020, with little difference between counties. From fall 2020 through January 2021, Facebook mobility levels for all counties decreased to slightly below baseline levels.

**Figure 7:**
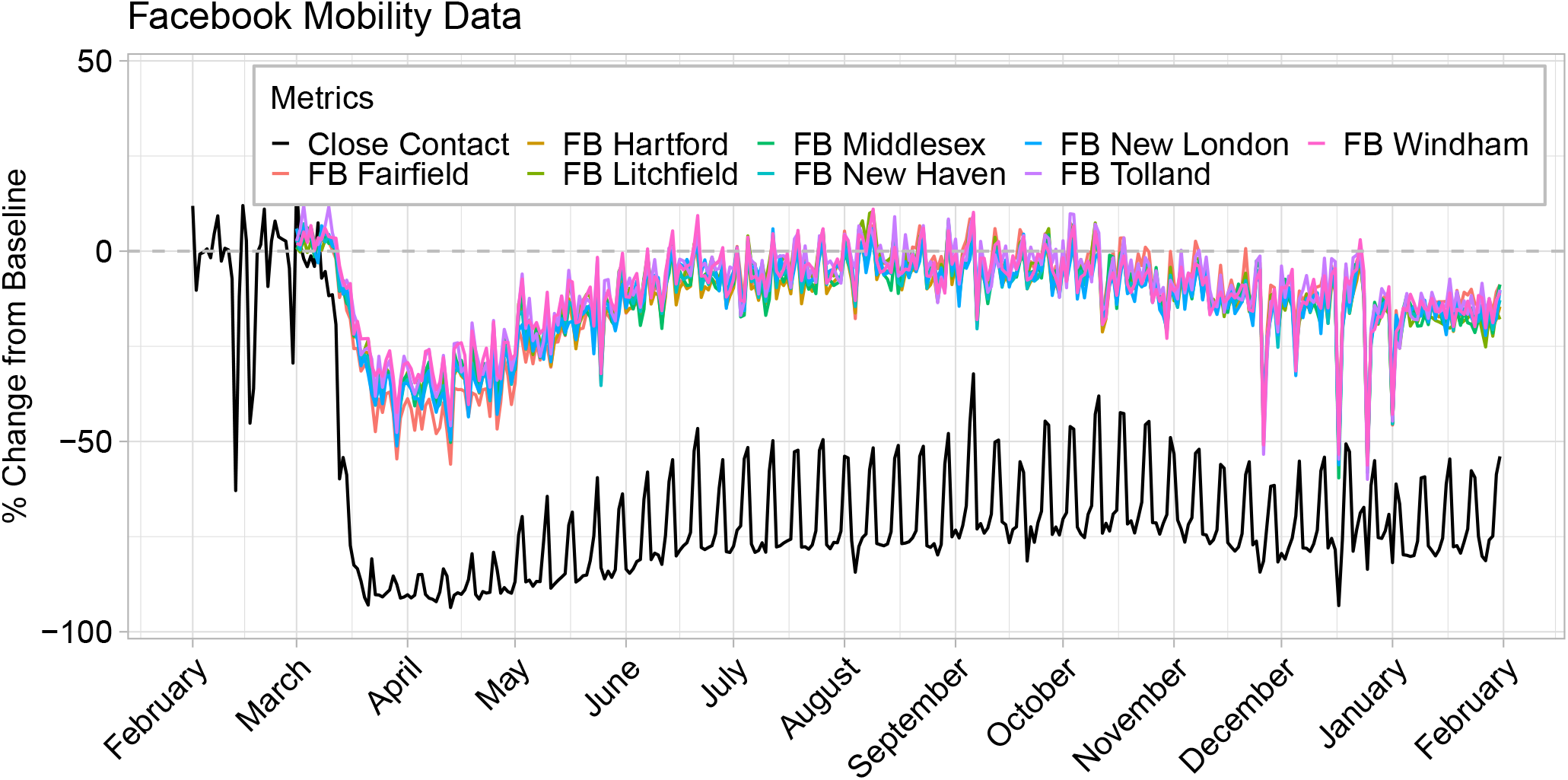
Comparison of Facebook (FB) mobility metrics to the contact rate described in this paper during February 1 – January 31, 2021.

Cuebiq county-level mobility data measures a 7 day rolling average of the median distance traveled in a day, and was available through November 1, 2020 ^1^. Figure 8 shows mobility data provided by Cuebiq with day-of-week median during February 2 – February 29, 2020 as the baseline. By July 2020, Cuebiq mobility levels returned to near baseline. Figure 9 shows Cuebiq’s metric for “contact”, when two or more devices are within 50 feet of each other within five minutes. Information about whether this metric takes spatial error (horizontal uncertainty) into account is not available. In July 2020, Cuebiq contact levels remained further below baseline than the Cuebiq mobility metric. The Cuebiq 50-foot contact metric was closer to baseline than our calculated contact rate during summer and fall 2020.

**Figure 8:**
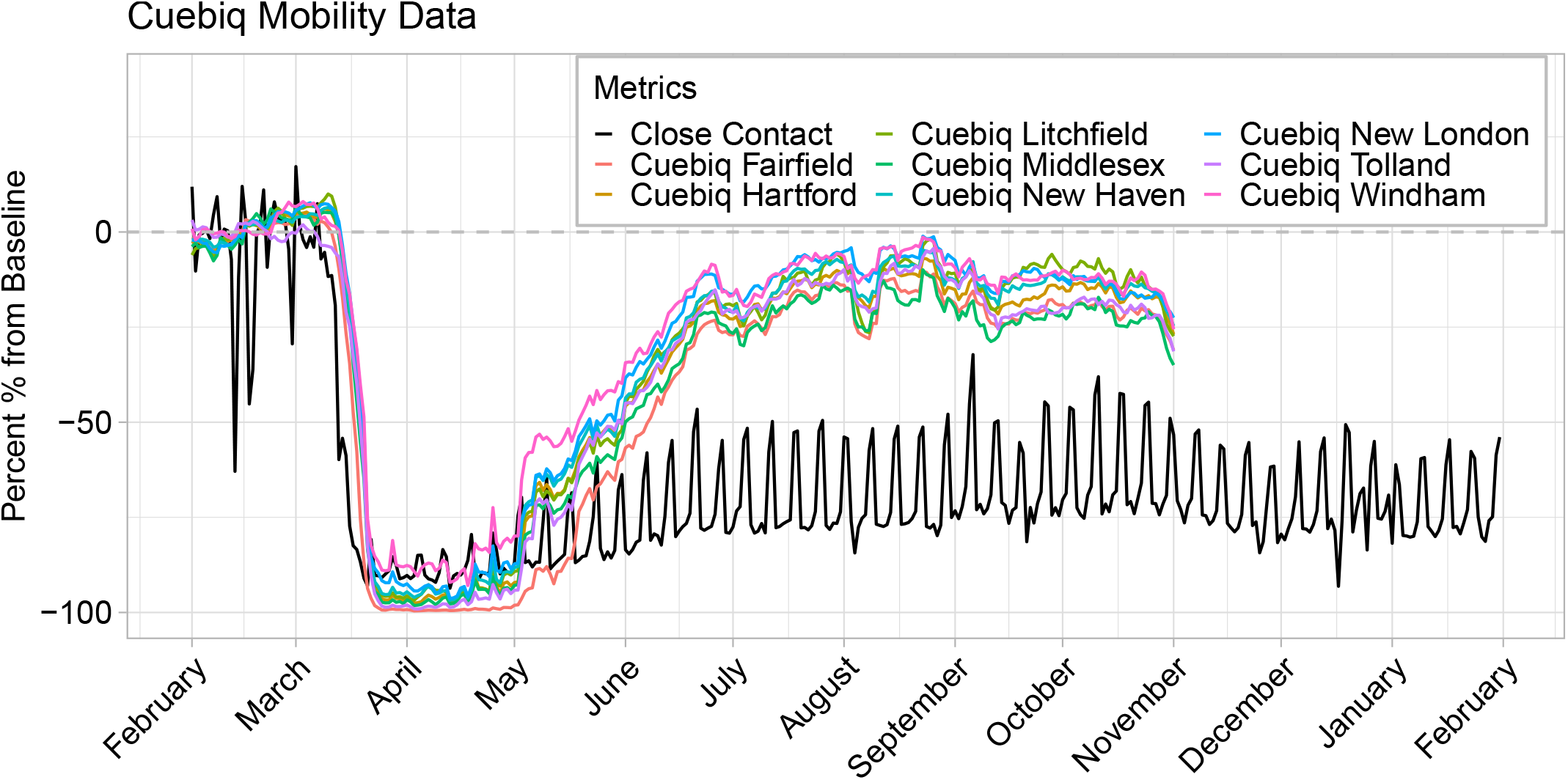
Comparison of Cuebiq mobility metrics to the contact rate described in this paper during February 1 – January 31, 2021. Cuebiq data available through November 1, 2020.

**Figure 9:**
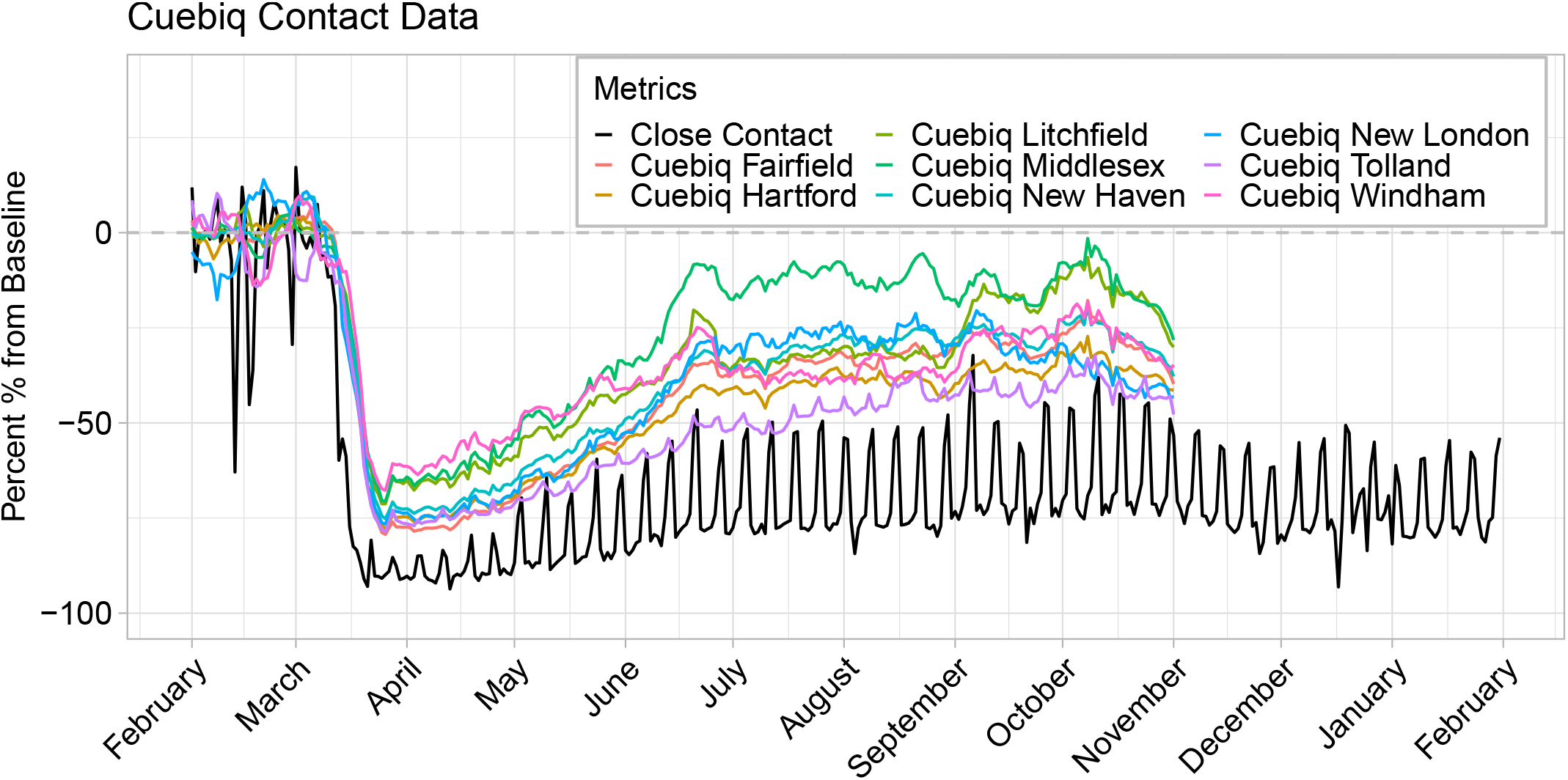
Comparison of the 50-foot Cuebiq contact metric to the contact rate described in this paper during February 1 – January 31, 2021. Cuebiq data available through November 1, 2020.

Finally, Descartes Labs state-level mobility data represents maximum distance devices have moved from the first reported location in a given day. Figure 10 shows the mobility metric provided by Descartes Labs with day-of-week median during February 17 – March 7, 2020 as the baseline. It was exceptional amongst the data sources in that mobility remained notably below baseline during March 2020 – January 2021. However, the percent decline in close contact was consistently larger than the observed percent decline in the Descartes Labs mobility metric.

**Figure 10:**
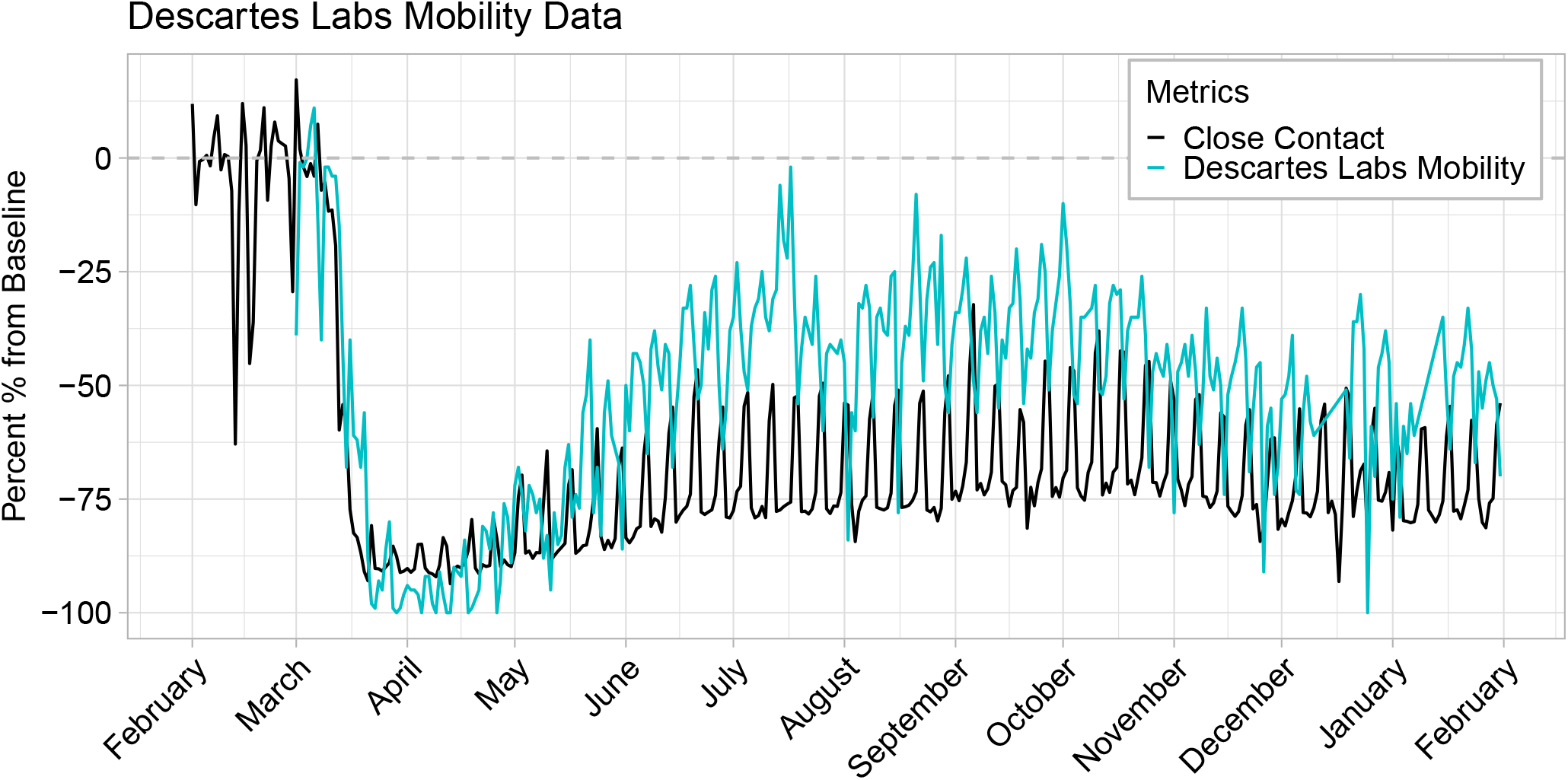
Comparison of Descartes Labs mobility metrics to the contact rate described in this paper during February 1 – January 31, 2021.

Every mobility metric we studied returns to baseline faster than the contact rate, though Descartes Labs’ mobility data provide the closest match to contact when presented on the percent change from February scale. In general, mobility metrics rebounded to February 2020 levels during summer 2020, and remained near this baseline or declined slightly through fall and winter 2020. Close contact, however, remained well below baseline levels through January 2021, suggesting that trends in mobility do not necessarily align with trends in close contact.

### 2.4 Space-time regression model results

In Table 2, we display the WAIC and posterior predictive deviance model comparison results. WAIC clearly favors the full spatio-temporal method even though the model is more complex than the two competitors as indicated by the effective number of parameters (p_WAIC_). This improvement in model fit however, overpowers this increased penalization term, leading to an improved overall balance. In terms of prediction, the full model is again preferred and seems to predict data with similar features to the observed data better than the competing models. Global Contact fits and predicts better than No Contact, suggesting that including contact information in some form within the model leads to improvements in both areas. Overall, these results show the importance of including prior day contact information in a flexible manner in order to explain variability in COVID-19 cases. Based on these findings, we further explore results from the full spatio-temporal model.

**Table 2:** Model fit (WAIC), complexity (p_WAIC_), and posterior predictive (ppd) deviance results from each of the competing methods.

| Method | WAIC | p<sub>WAIC</sub> | PP Deviance |
| --- | --- | --- | --- |
| Full Spatiotemporal | **187,849.30** | 612.45 | **68,177.60** |
| Global Contact | 188,562.30 | 334.83 | 69,360.68 |
| No Contact | 188,680.40 | 328.88 | 69,535.07 |

In Table 3, we display posterior inference for the non-contact regression parameters (*β*_*j*_), exponentiated for a relative risk interpretation. Results suggest that towns with increased population have more cases on average, and that towns with higher median household income and a larger proportion of residents that are non-Hispanic White and ≥ 65 years have fewer cases. None of the other included spatially-varying covariates had 95% credible intervals that excluded one.

**Table 3:** Regression parameter posterior inference (posterior means and quantile-based 95% credible intervals) from the full spatio-temporal model. (a) 10,000 increase; (b) 10% increase.

| Variable | Relative Risk |  |
| --- | --- | --- |
|  | Mean | 95% CrI |
| Population<sup>a</sup> | 1.34 | (1.24, 1.44) |
| Median Household Income (USD)<sup>a</sup> | 0.94 | (0.88, 0.99) |
| % Poverty<sup>b</sup> | 0.88 | (0.66, 1.15) |
| % White, Non-Hispanic<sup>b</sup> | 0.89 | (0.81, 0.99) |
| % Some College Education<sup>b</sup> | 0.95 | (0.67, 1.31) |
| % $\geq 65$ Years<sup>b</sup> | 0.77 | (0.58, 0.98) |

In Figure 11, we show the estimated global lag parameters, ***θ***, and 95% credible intervals across the different lag days from the full spatio-temporal version of the model. The figure suggests that increased contact on prior days 3-7 is associated with increased current day COVID-19 cases.

**Figure 11:**
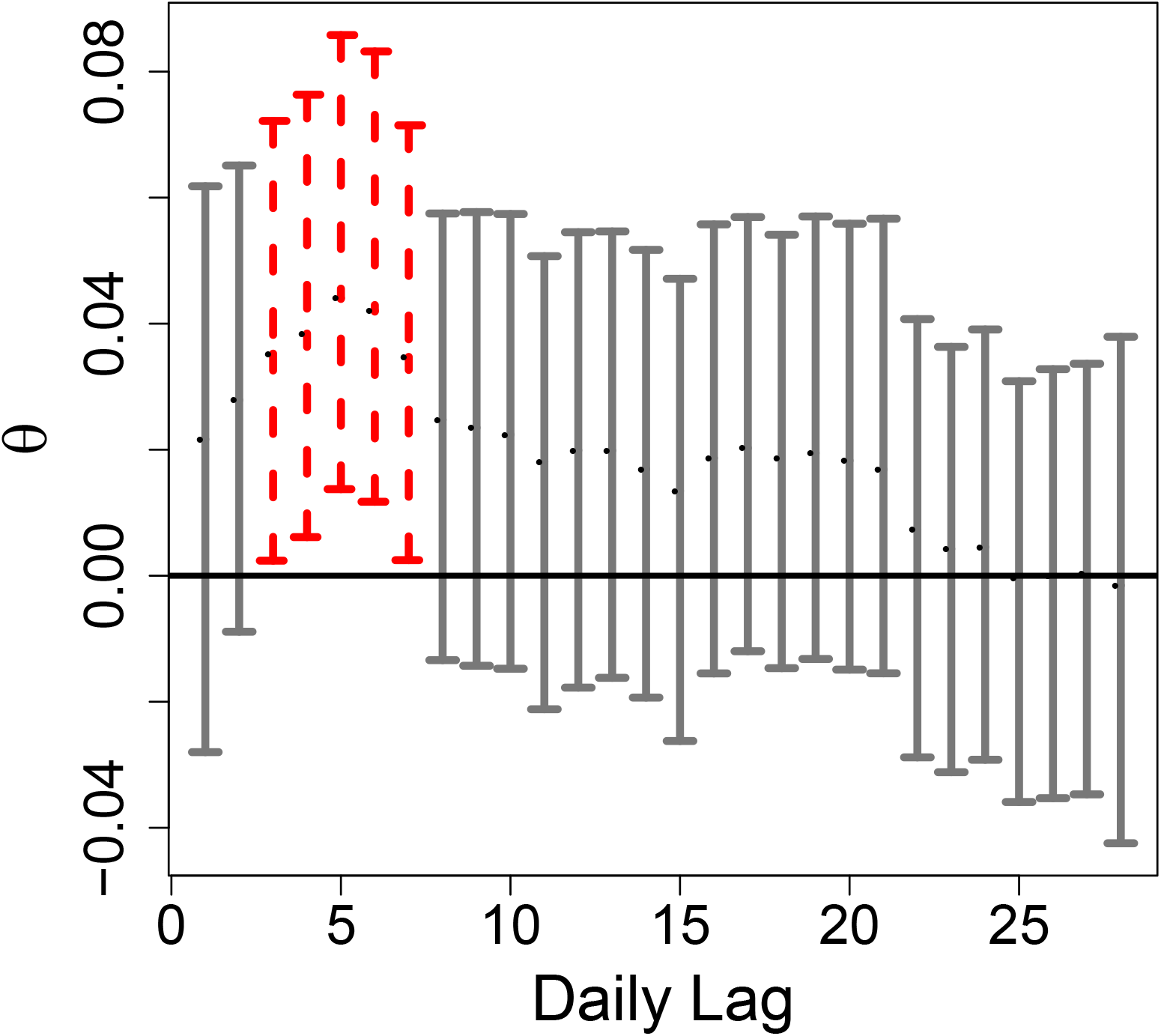
Estimated daily lag regression parameters (and 95% credible intervals) describing associations between contact and current day COVID-19 cases. Red bars indicate that the credible intervals exclude zero.

### 2.5 Transmission model predicted cases and estimated infections for each town

Figure 12 through 33 show contact, projected infections, cases, and cumulative incidence for each of the 169 towns in Connecticut.

**Figure 12:**
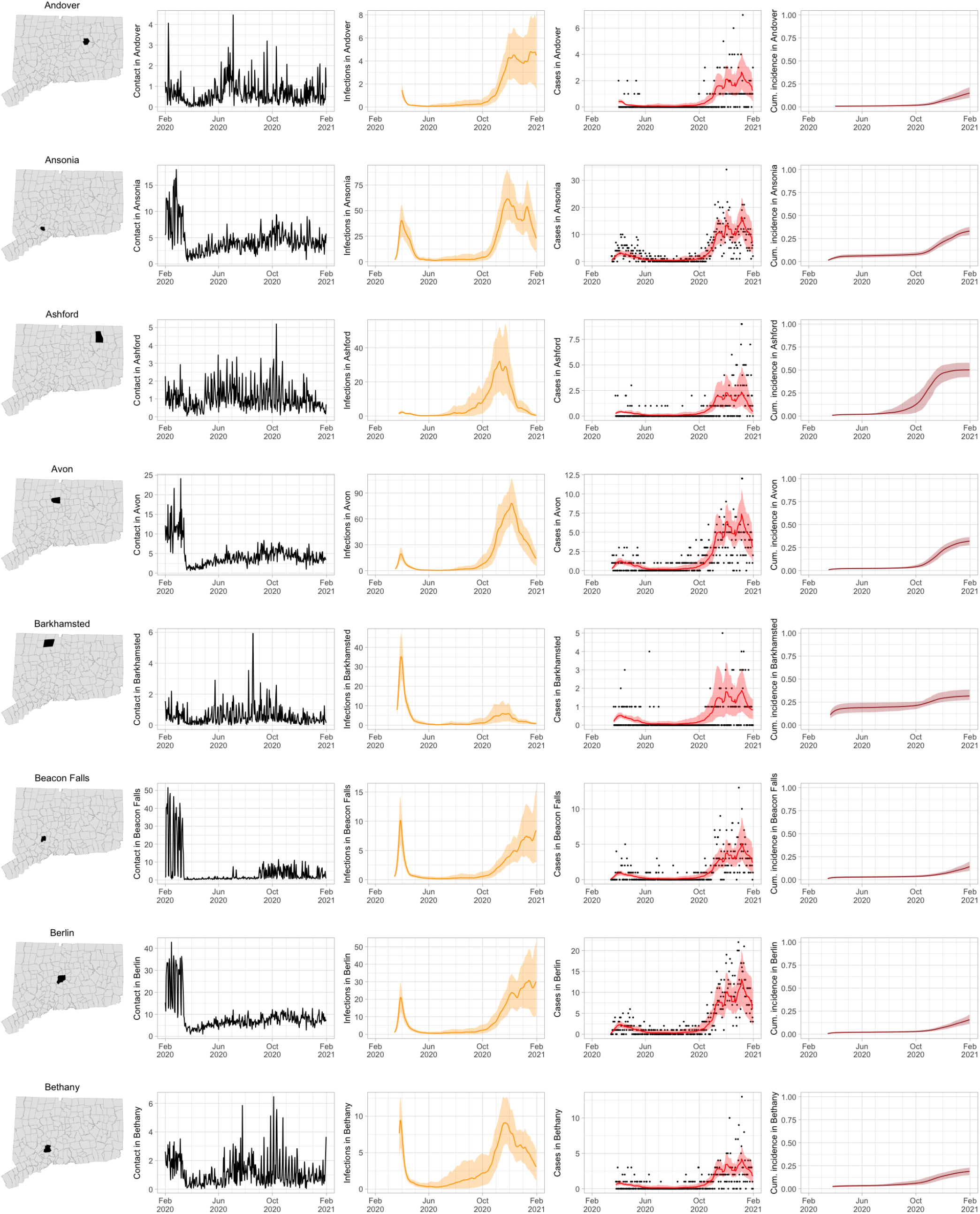
Contact for Connecticut towns and fitted SEIR model predictions with 95% uncertainty intervals.

**Figure 13:**
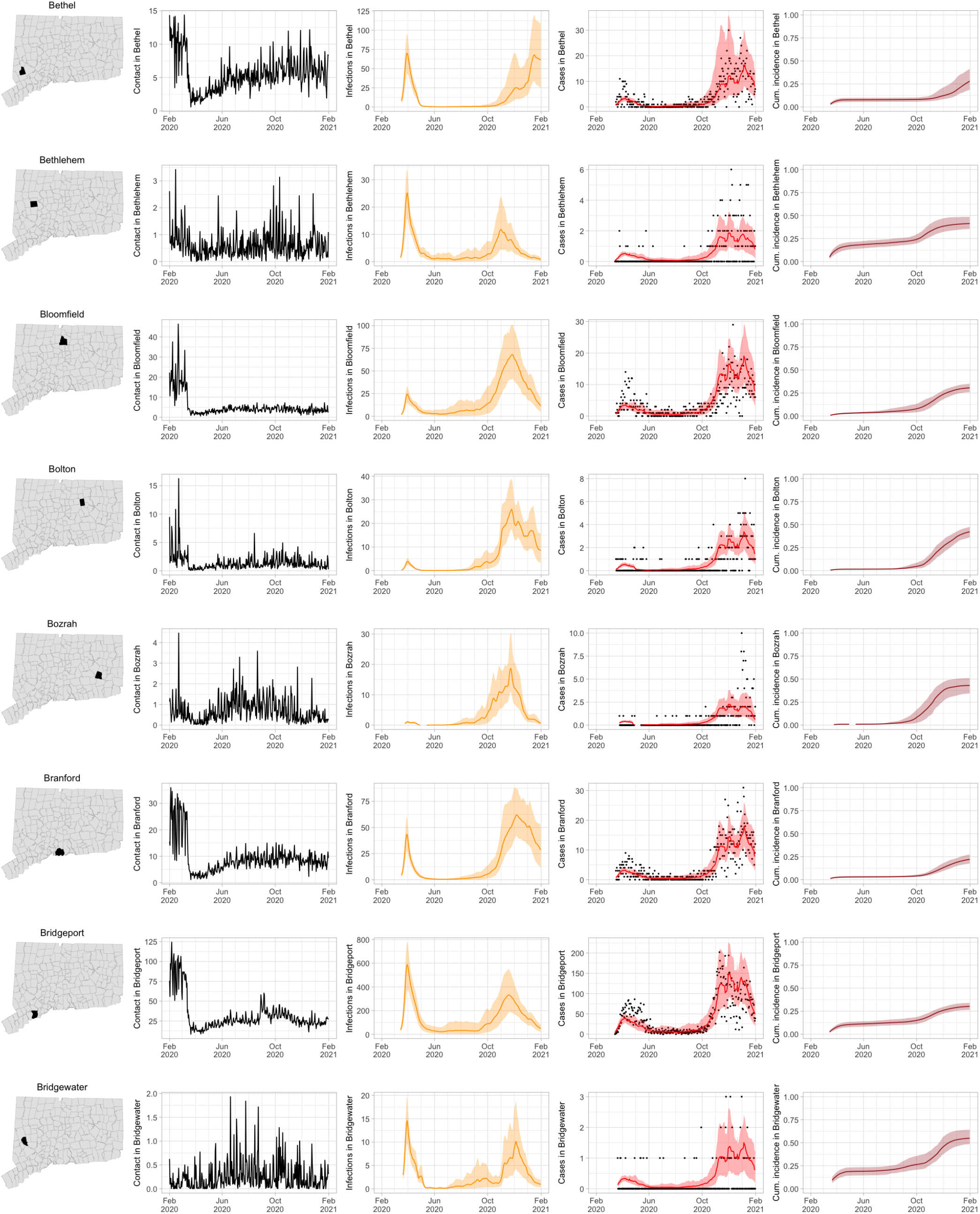
Contact for Connecticut towns and fitted SEIR model predictions with 95% uncertainty intervals.

**Figure 14:**
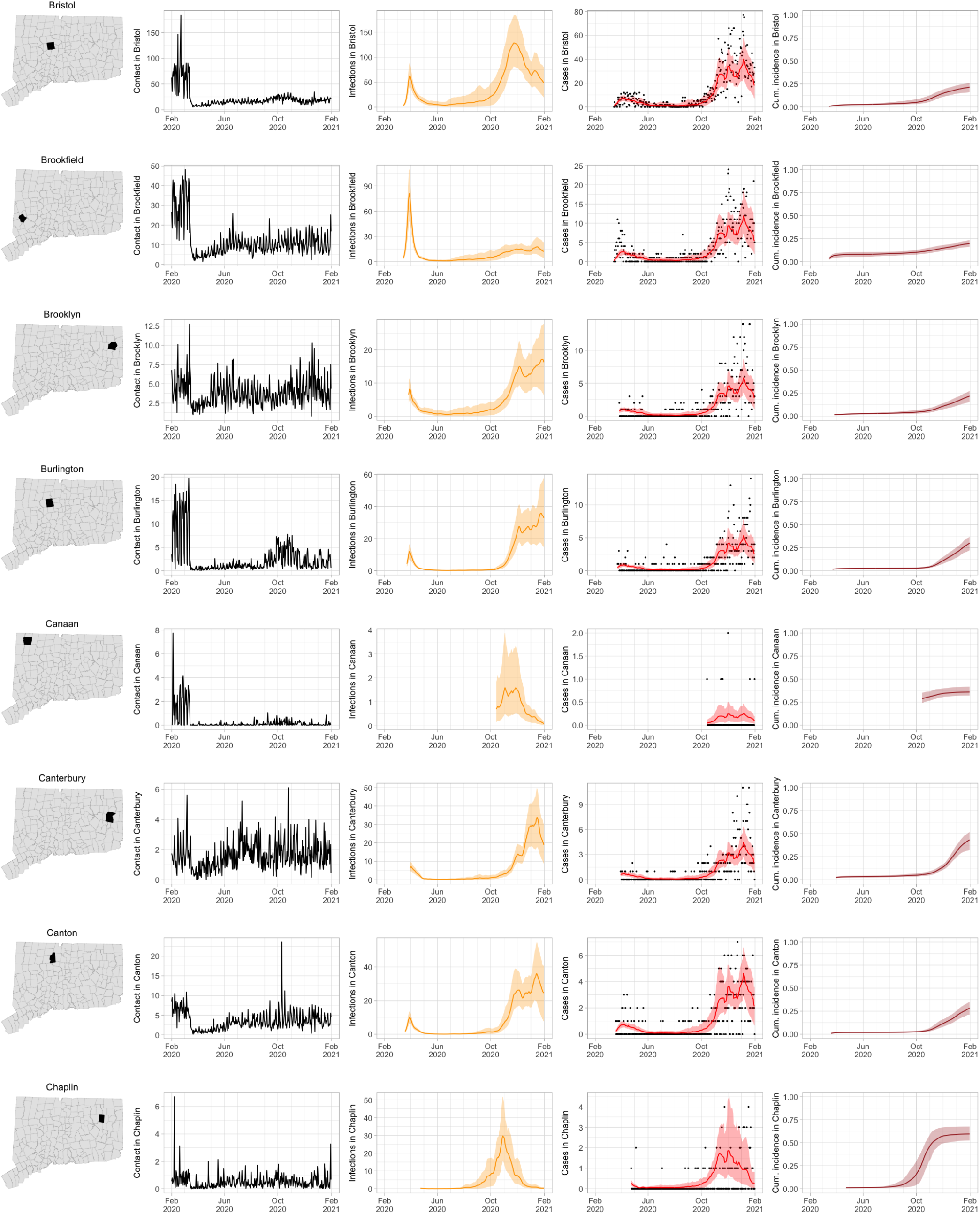
Contact for Connecticut towns and fitted SEIR model predictions with 95% uncertainty intervals.

**Figure 15:**
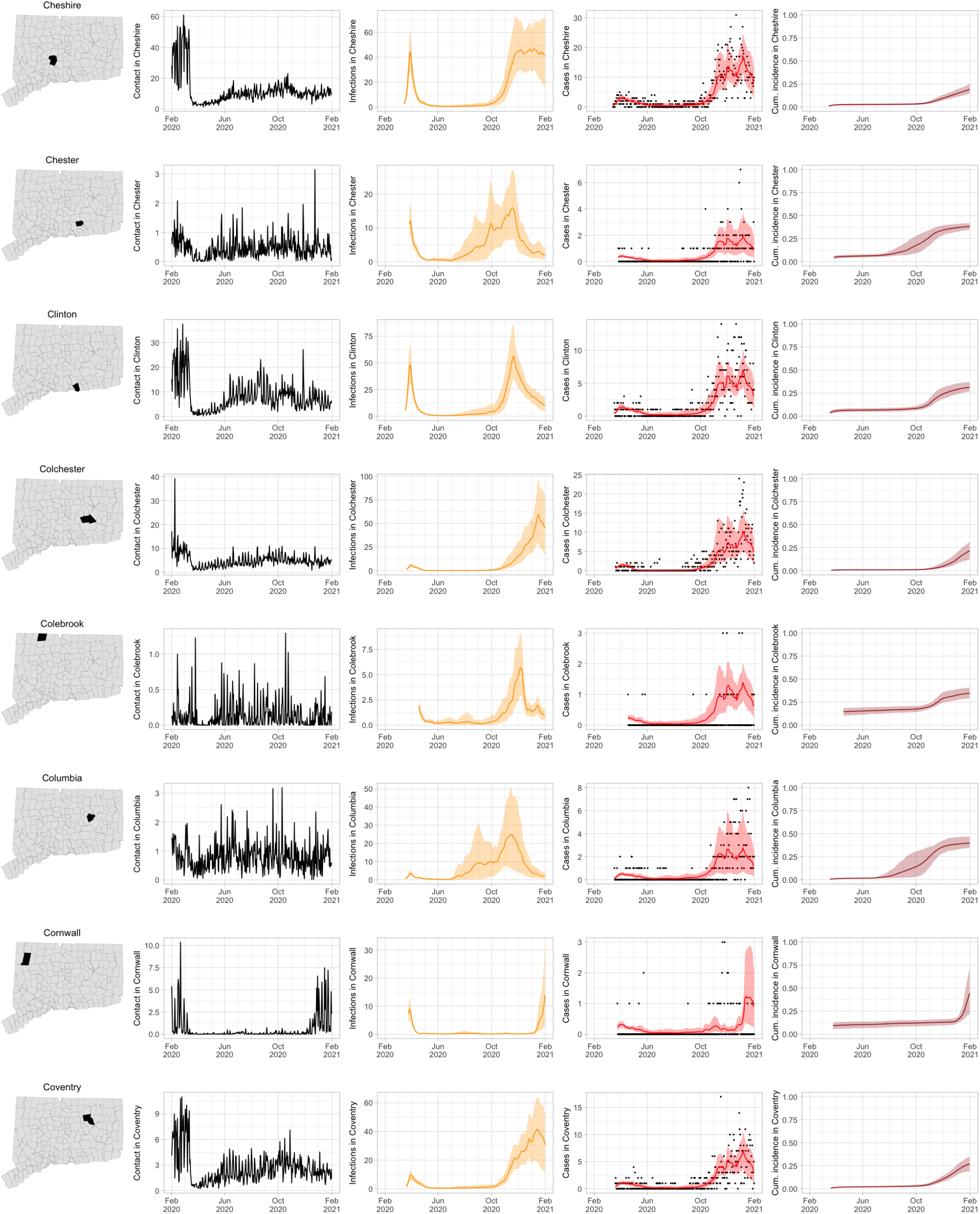
Contact for Connecticut towns and fitted SEIR model predictions with 95% uncertainty intervals.

**Figure 16:**
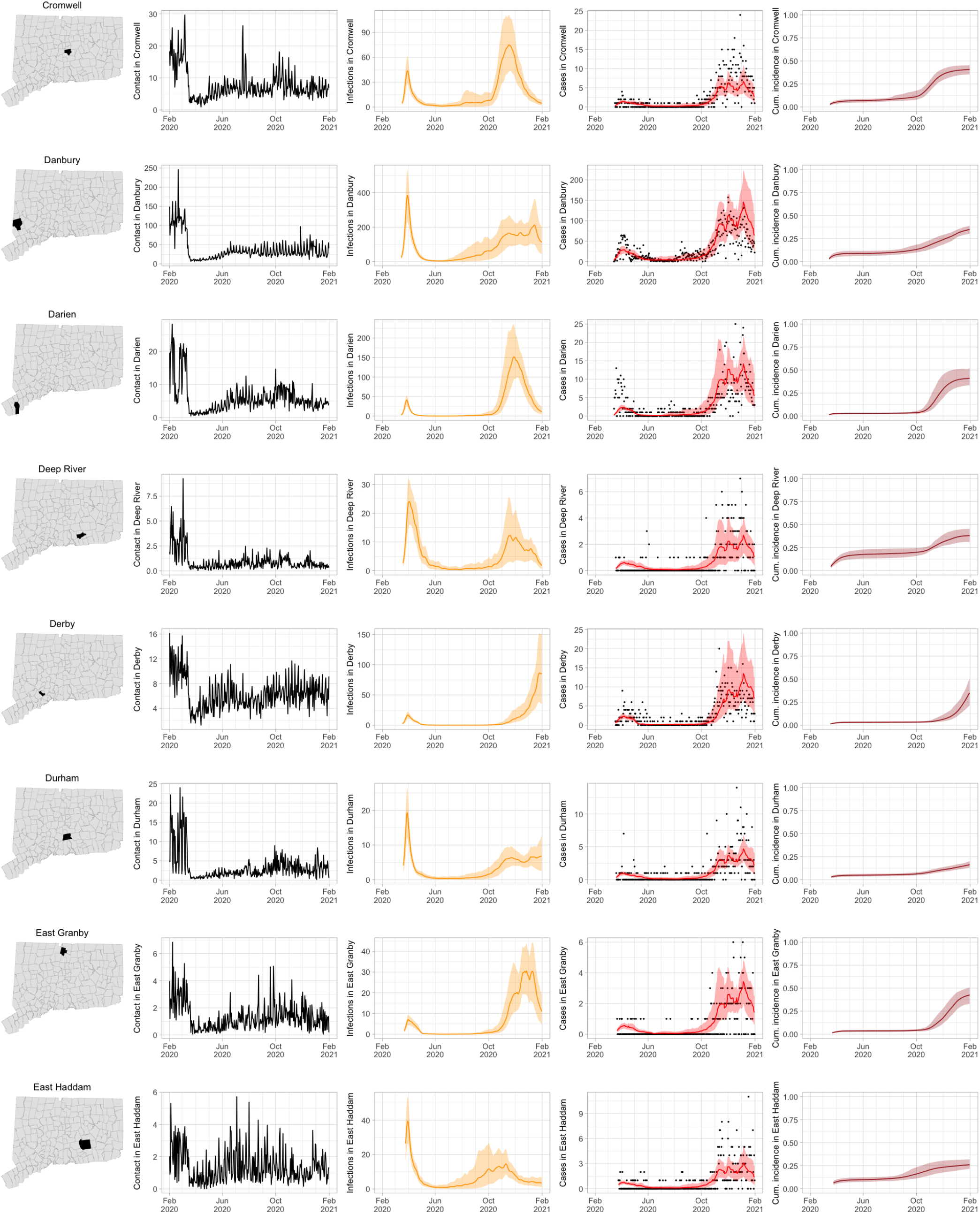
Contact for Connecticut towns and fitted SEIR model predictions with 95% uncertainty intervals.

**Figure 17:**
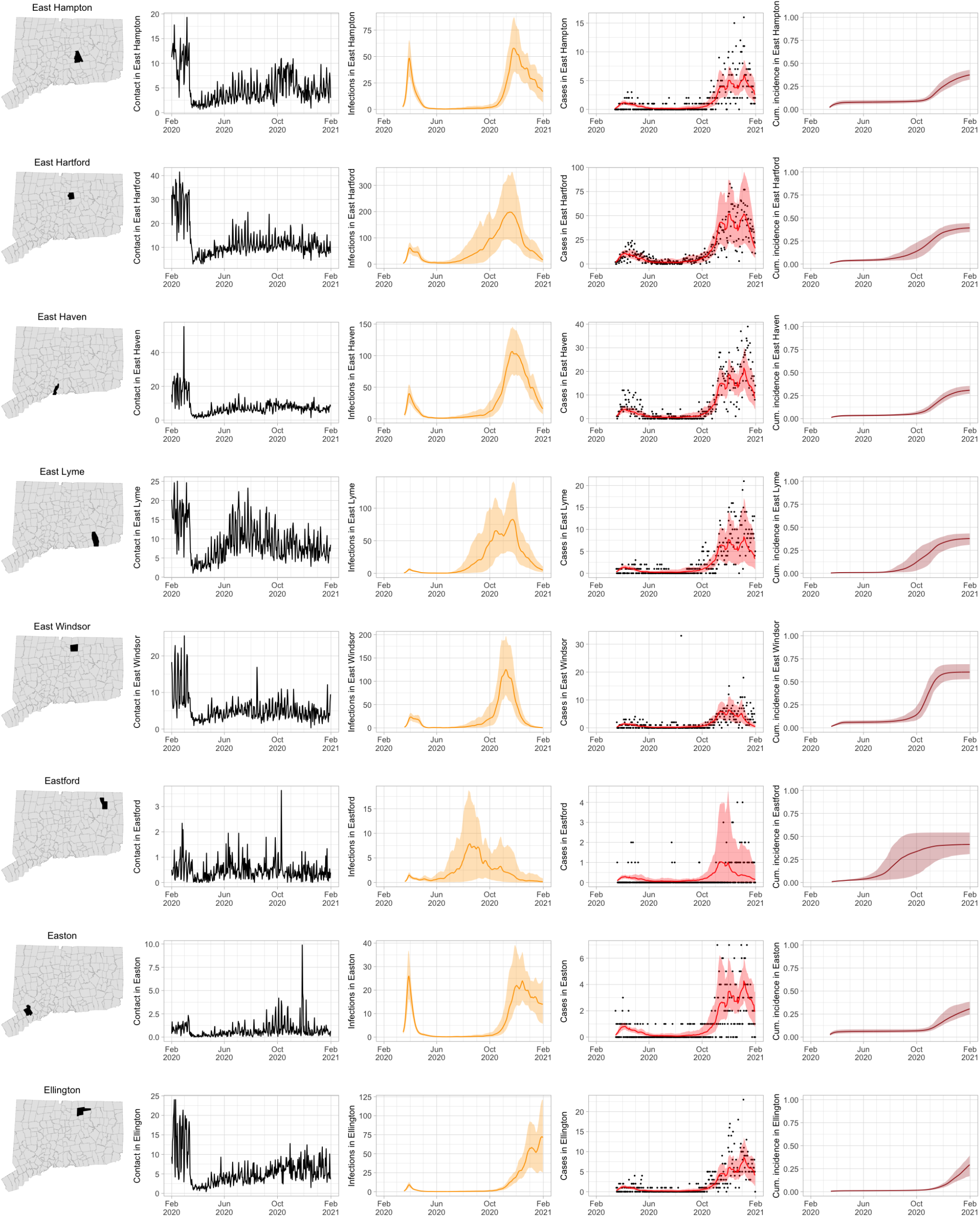
Contact for Connecticut towns and fitted SEIR model predictions with 95% uncertainty intervals.

**Figure 18:**
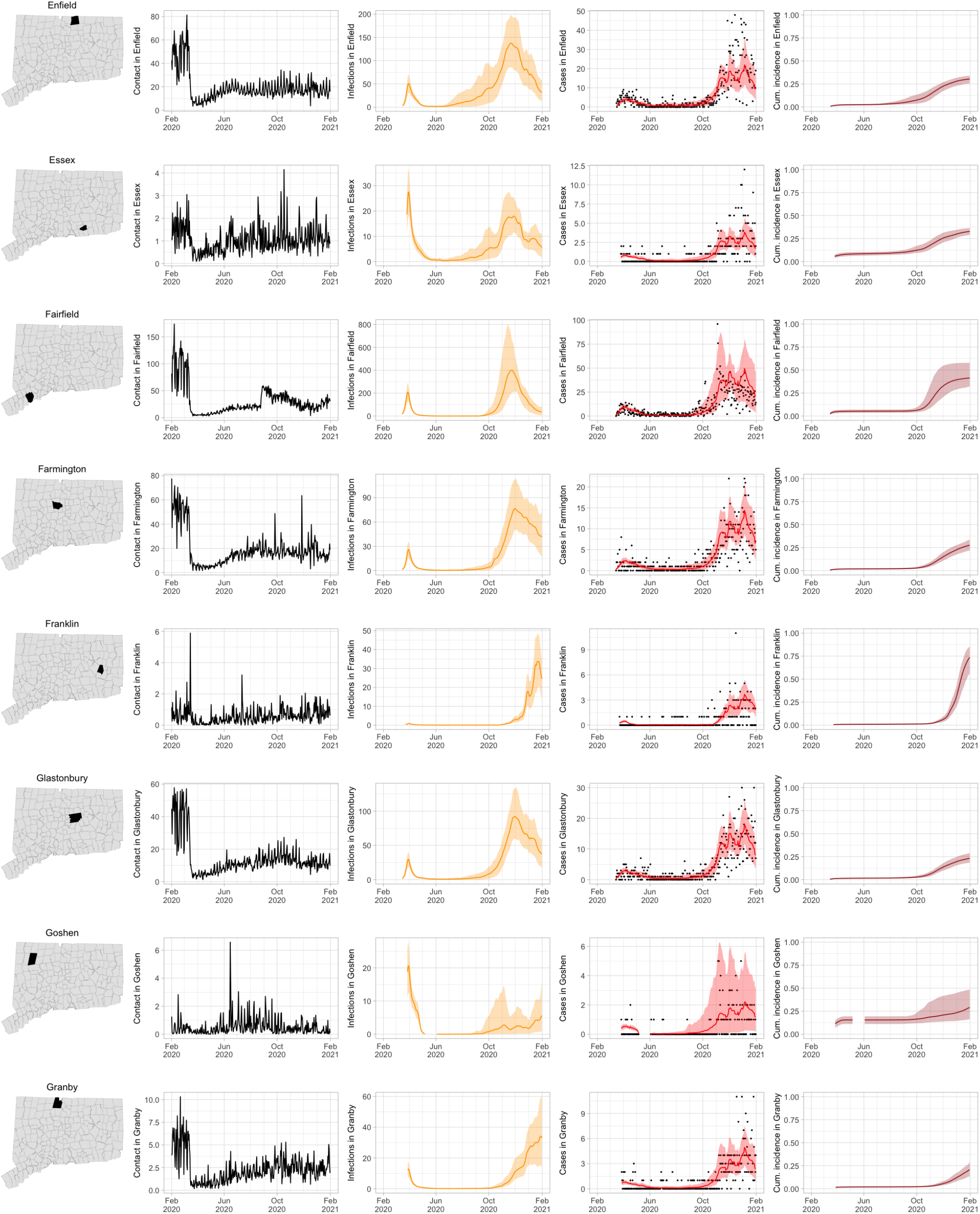
Contact for Connecticut towns and fitted SEIR model predictions with 95% uncertainty intervals.

**Figure 19:**
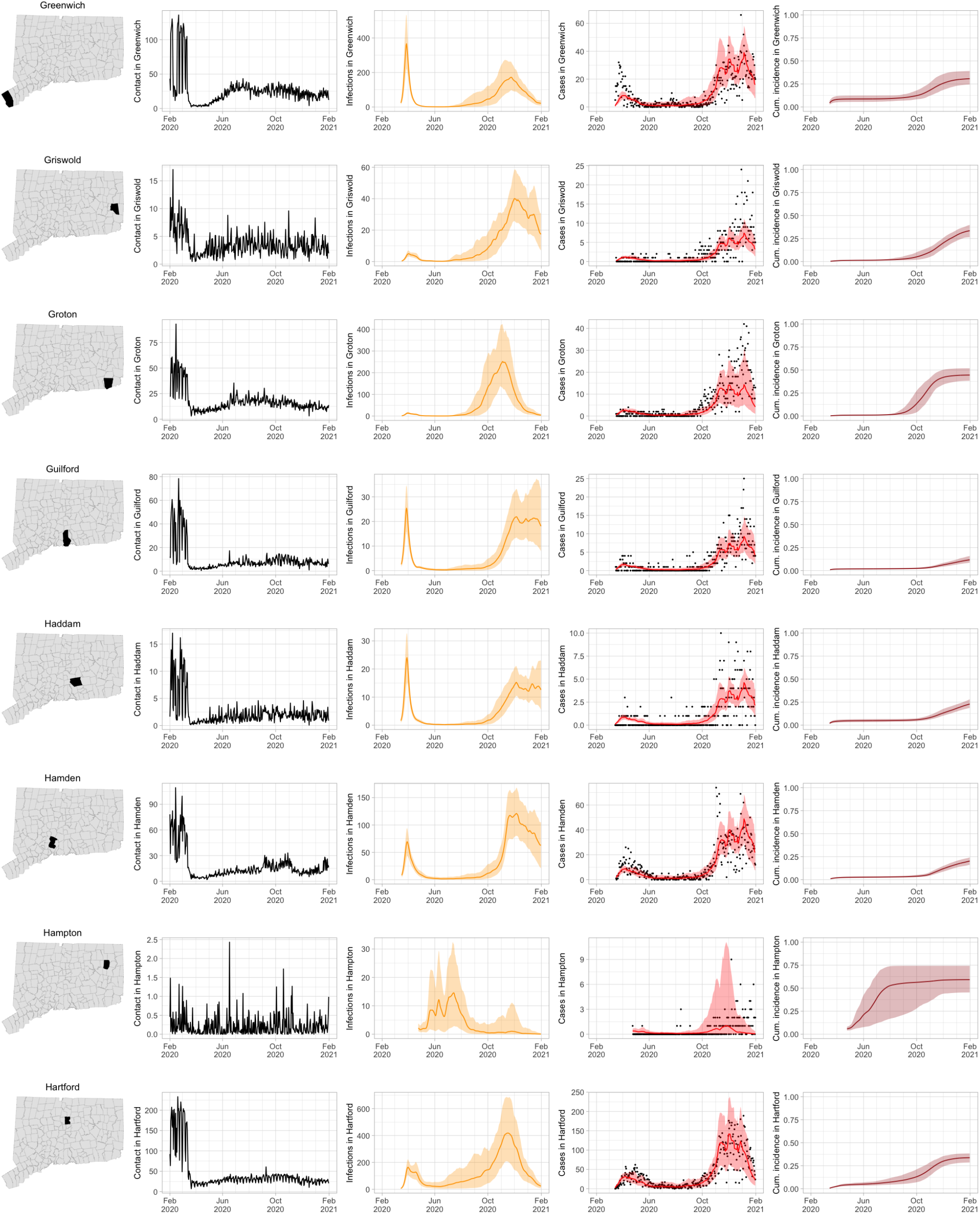
Contact for Connecticut towns and fitted SEIR model predictions with 95% uncertainty intervals.

**Figure 20:**
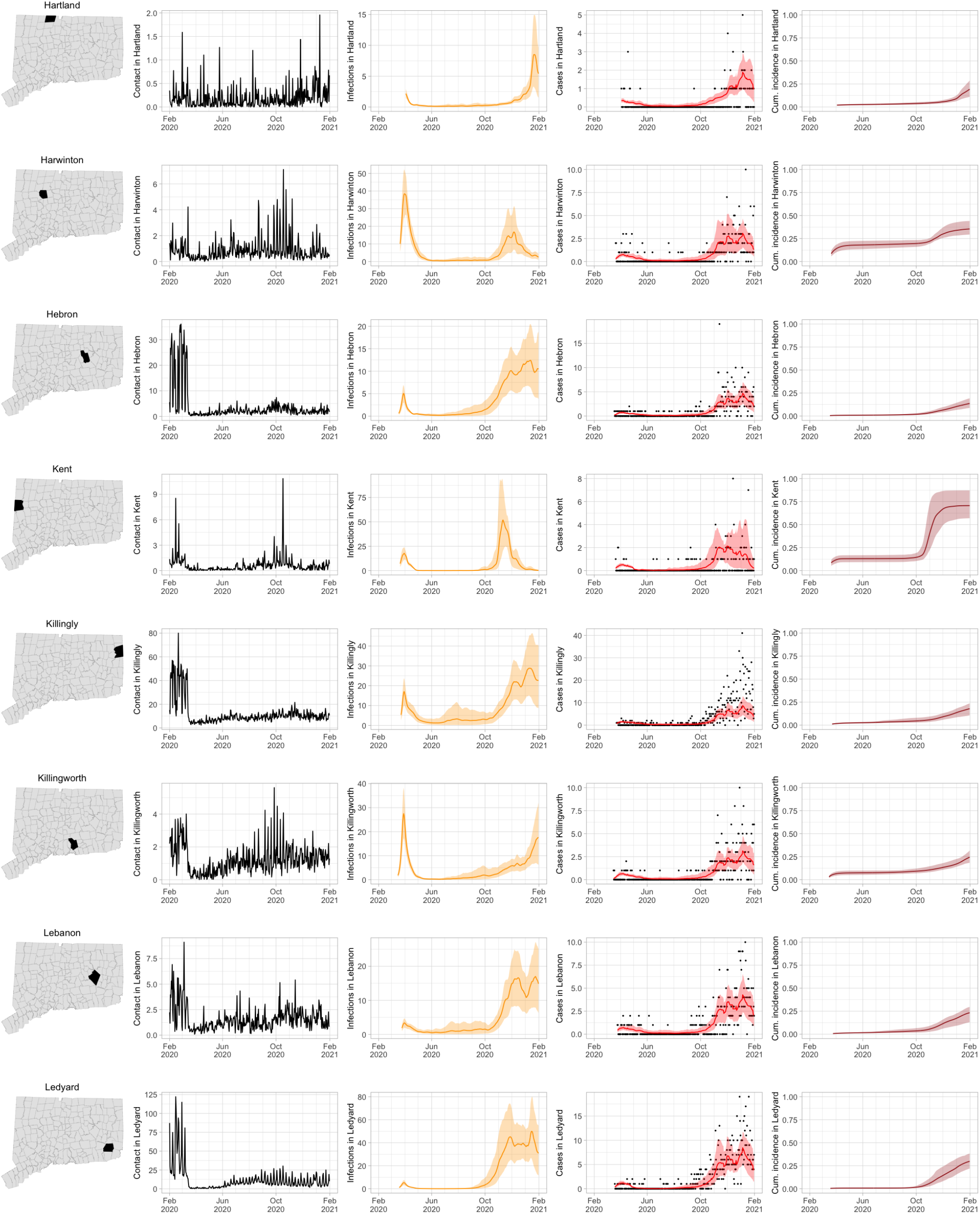
Contact for Connecticut towns and fitted SEIR model predictions with 95% uncertainty intervals.

**Figure 21:**
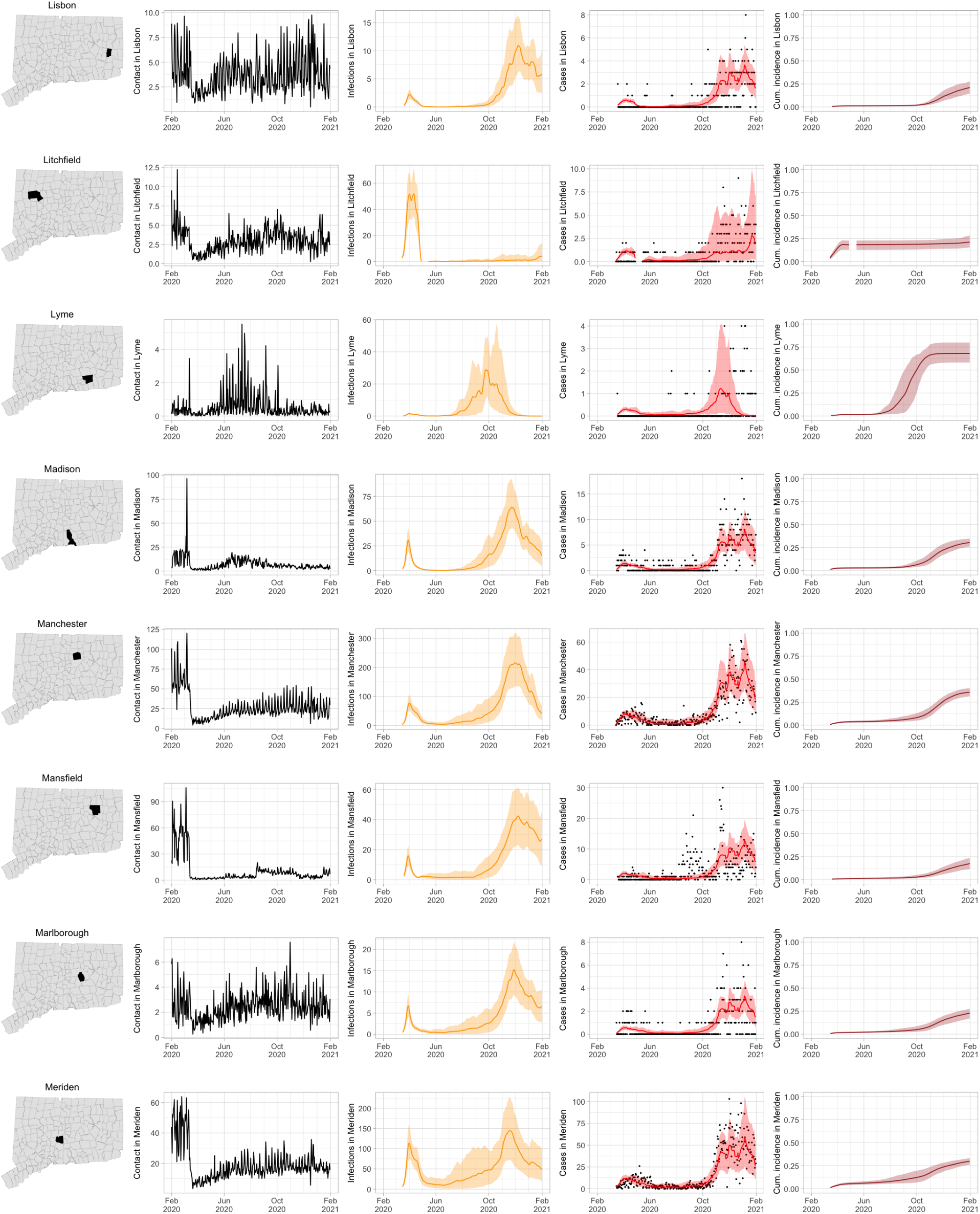
Contact for Connecticut towns and fitted SEIR model predictions with 95% uncertainty intervals.

**Figure 22:**
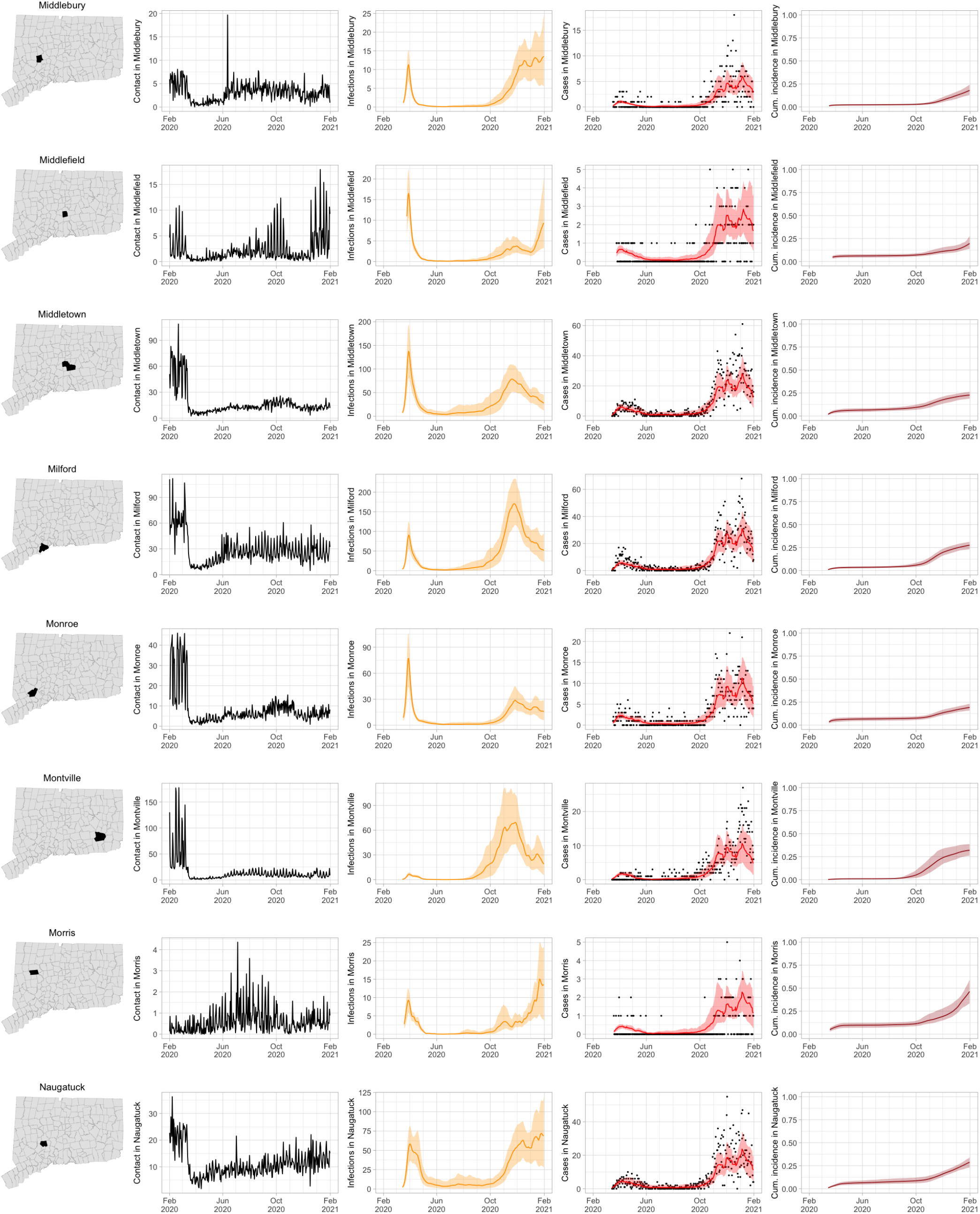
Contact for Connecticut towns and fitted SEIR model predictions with 95% uncertainty intervals.

**Figure 23:**
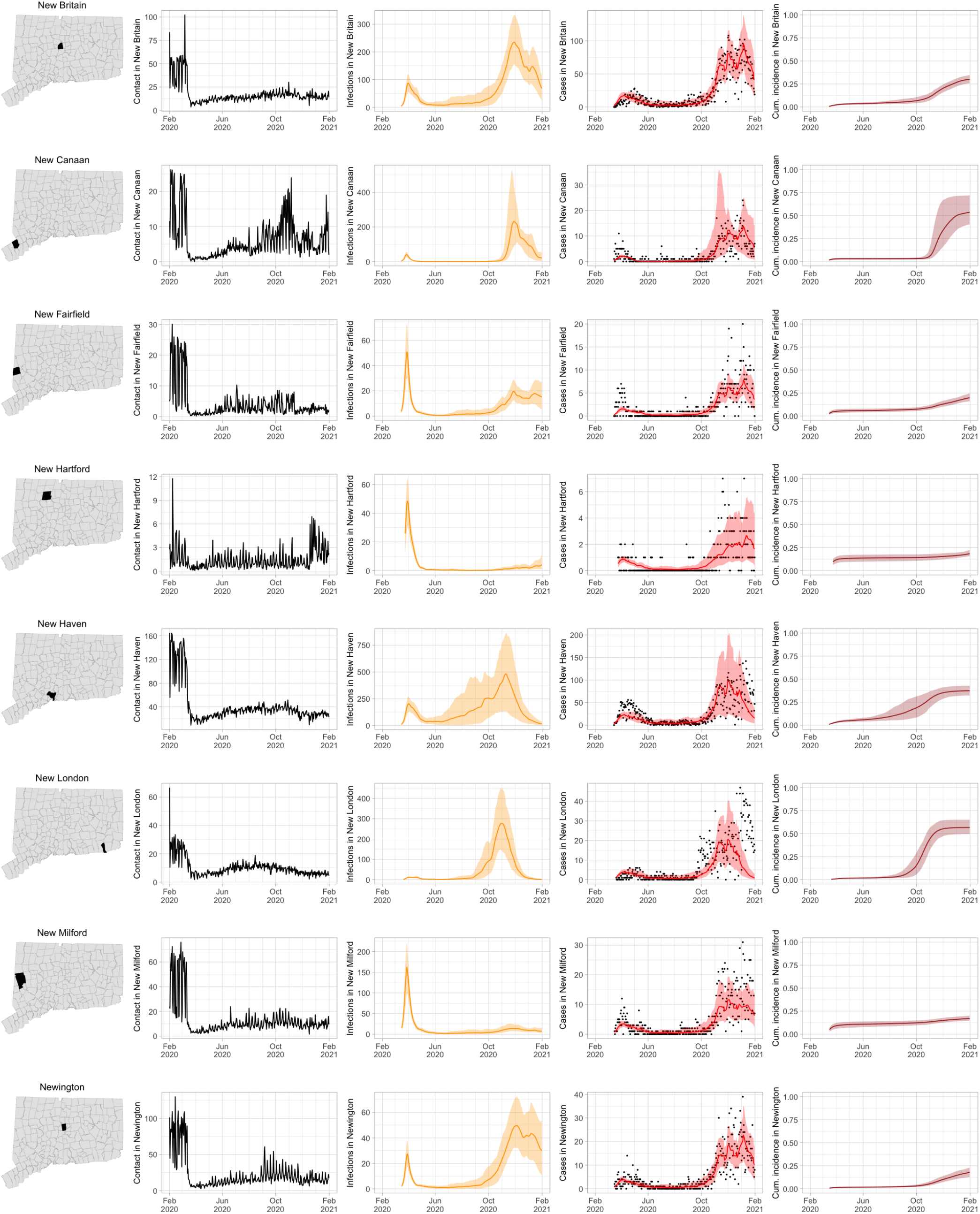
Contact for Connecticut towns and fitted SEIR model predictions with 95% uncertainty intervals.

**Figure 24:**
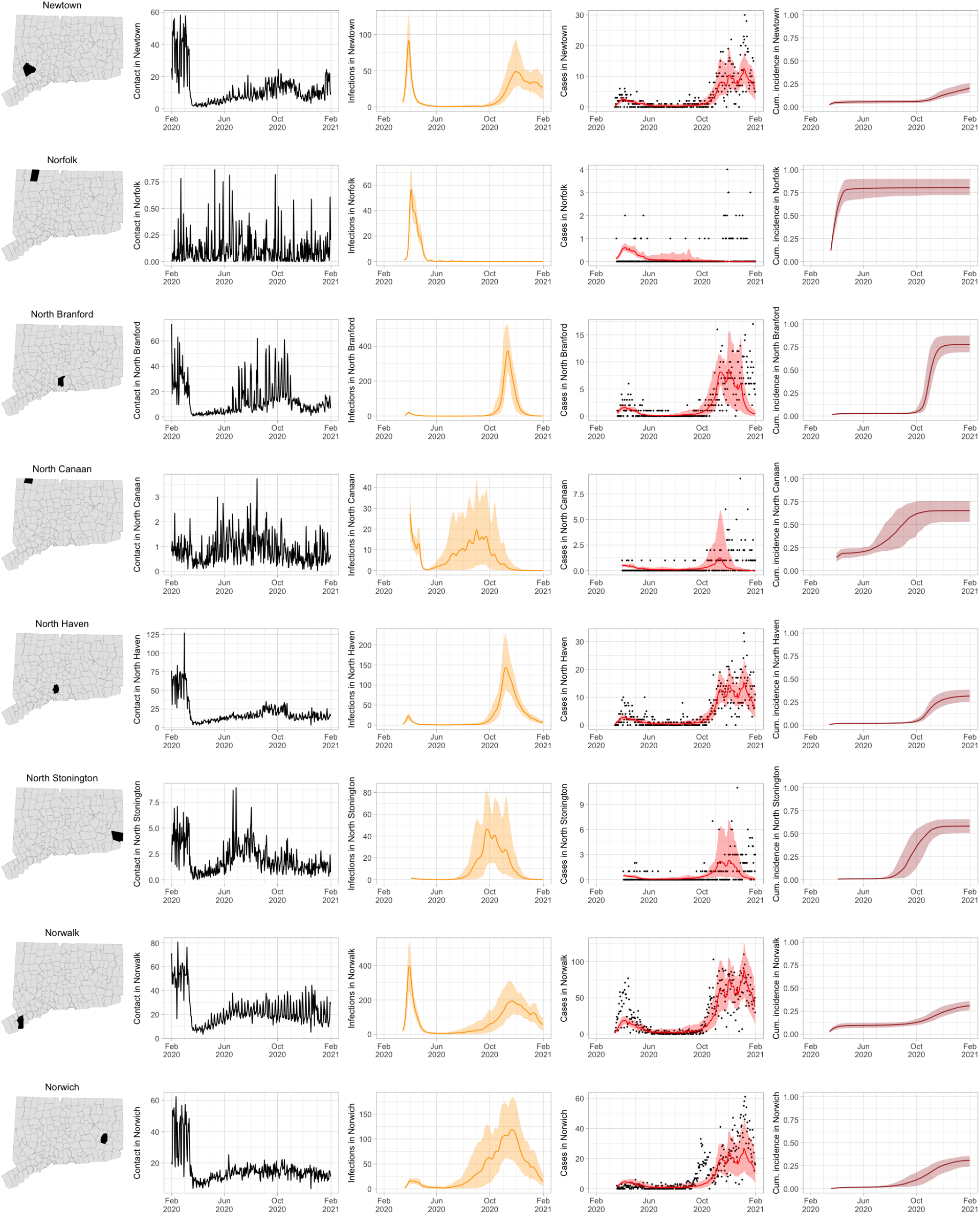
Contact for Connecticut towns and fitted SEIR model predictions with 95% uncertainty intervals.

**Figure 25:**
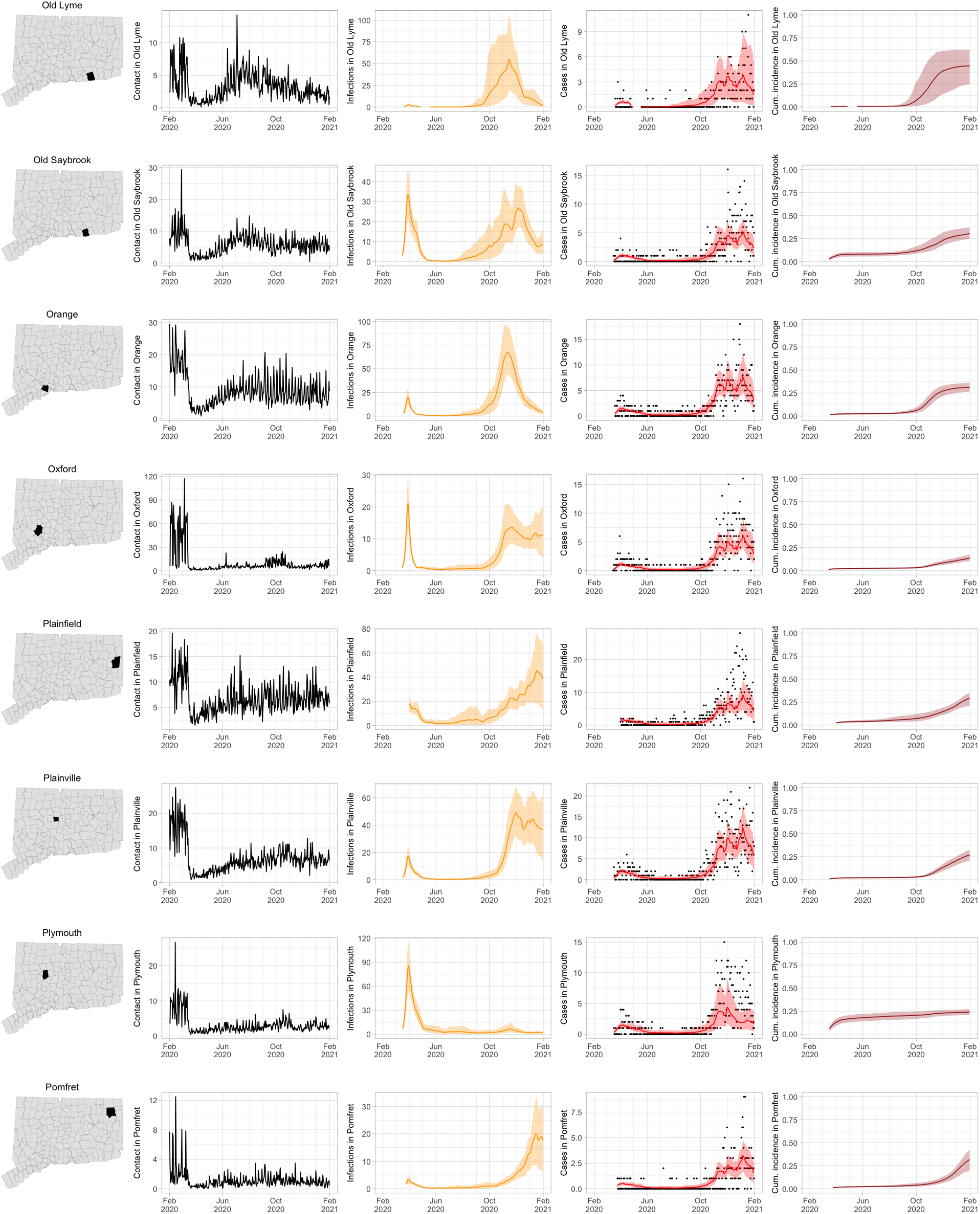
Contact for Connecticut towns and fitted SEIR model predictions with 95% uncertainty intervals.

**Figure 26:**
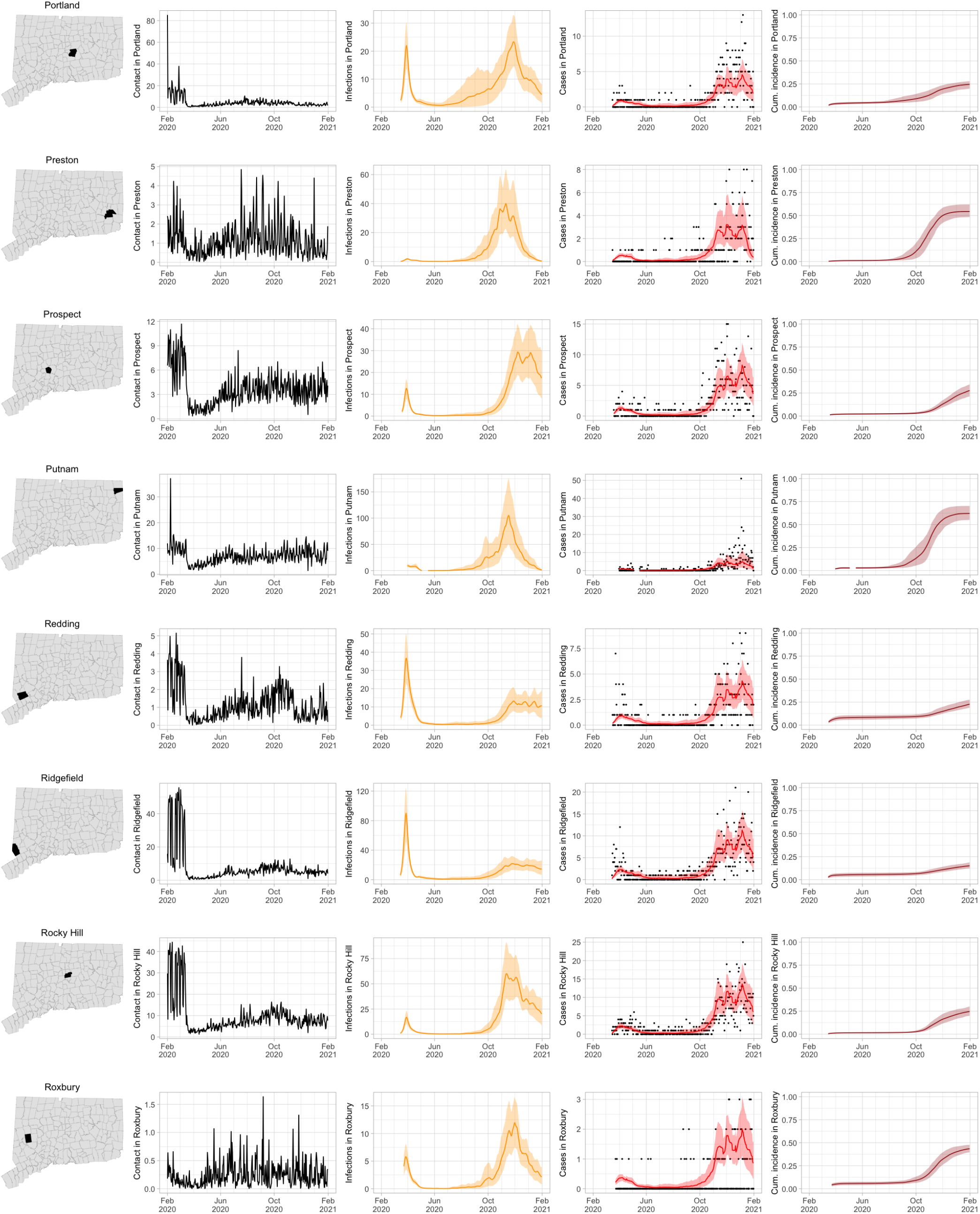
Contact for Connecticut towns and fitted SEIR model predictions with 95% uncertainty intervals.

**Figure 27:**
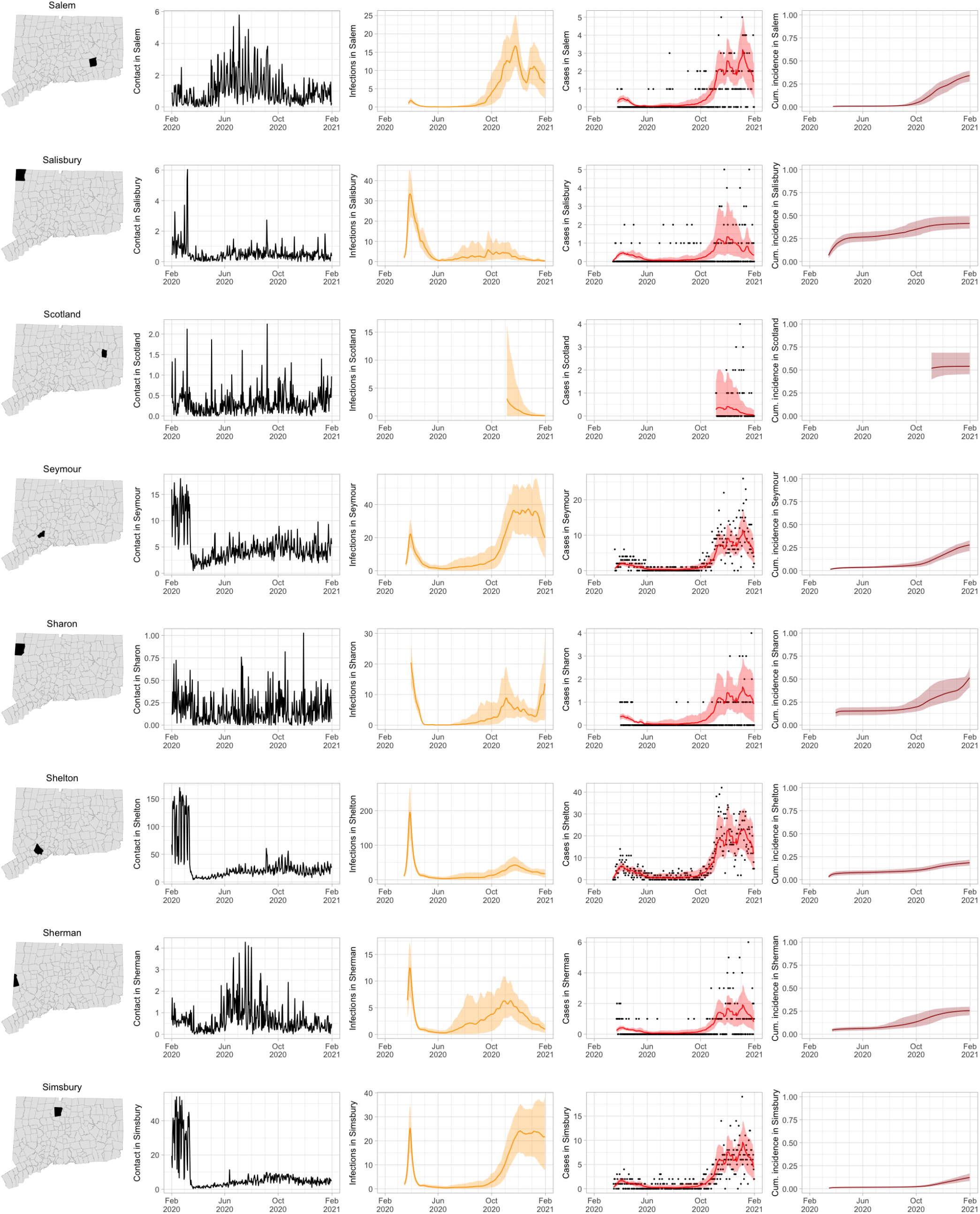
Contact for Connecticut towns and fitted SEIR model predictions with 95% uncertainty intervals.

**Figure 28:**
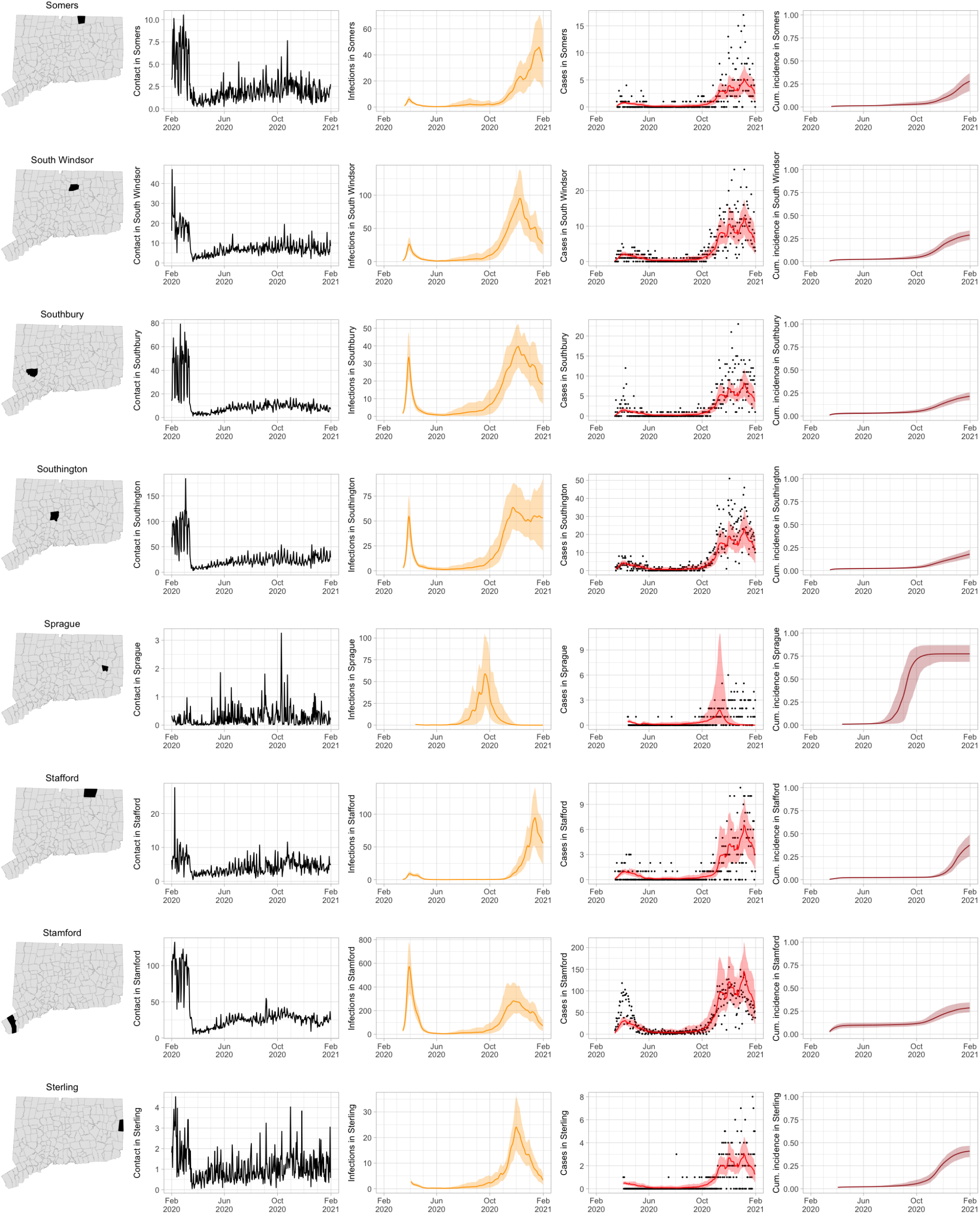
Contact for Connecticut towns and fitted SEIR model predictions with 95% uncertainty intervals.

**Figure 29:**
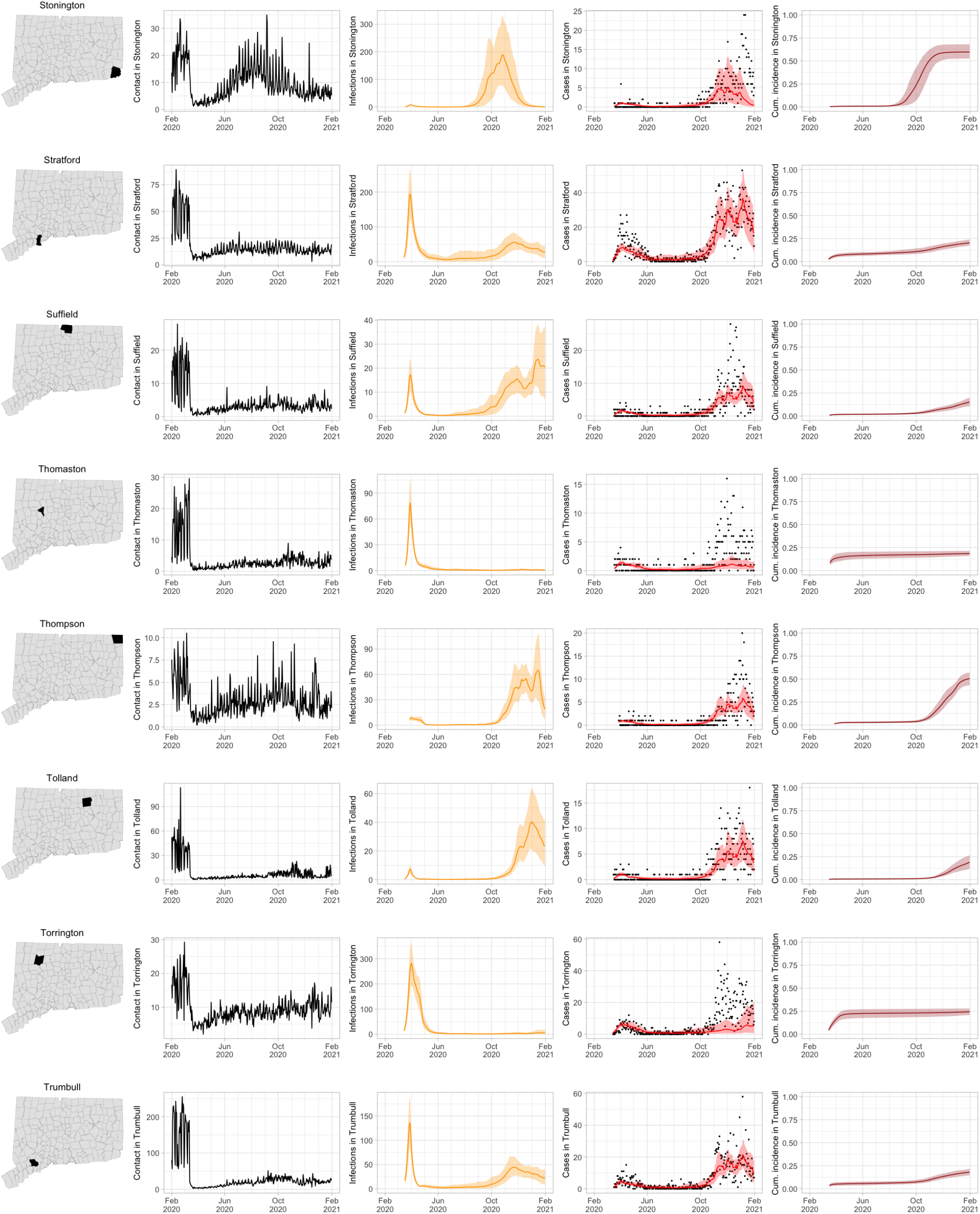
Contact for Connecticut towns and fitted SEIR model predictions with 95% uncertainty intervals.

**Figure 30:**
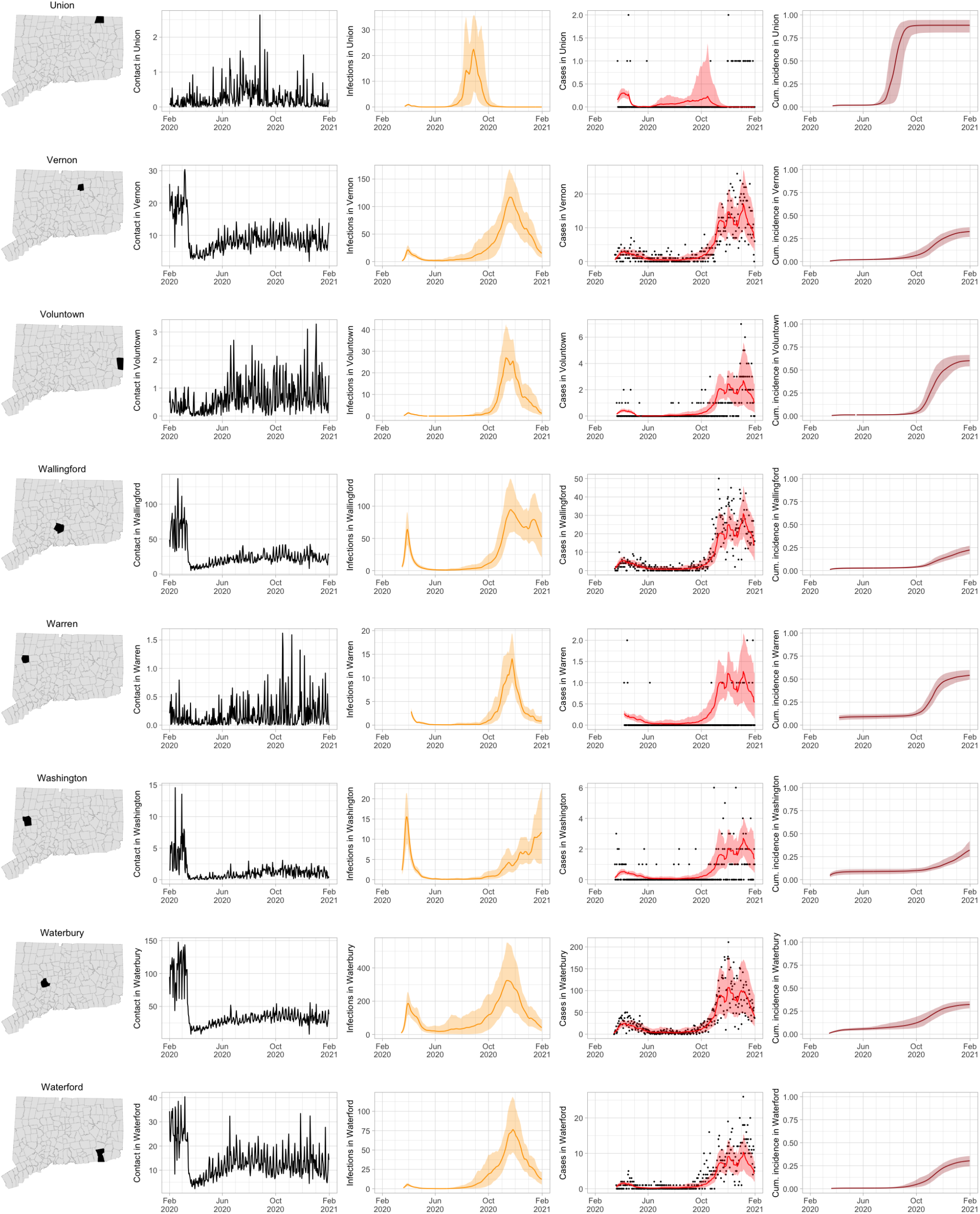
Contact for Connecticut towns and fitted SEIR model predictions with 95% uncertainty intervals.

**Figure 31:**
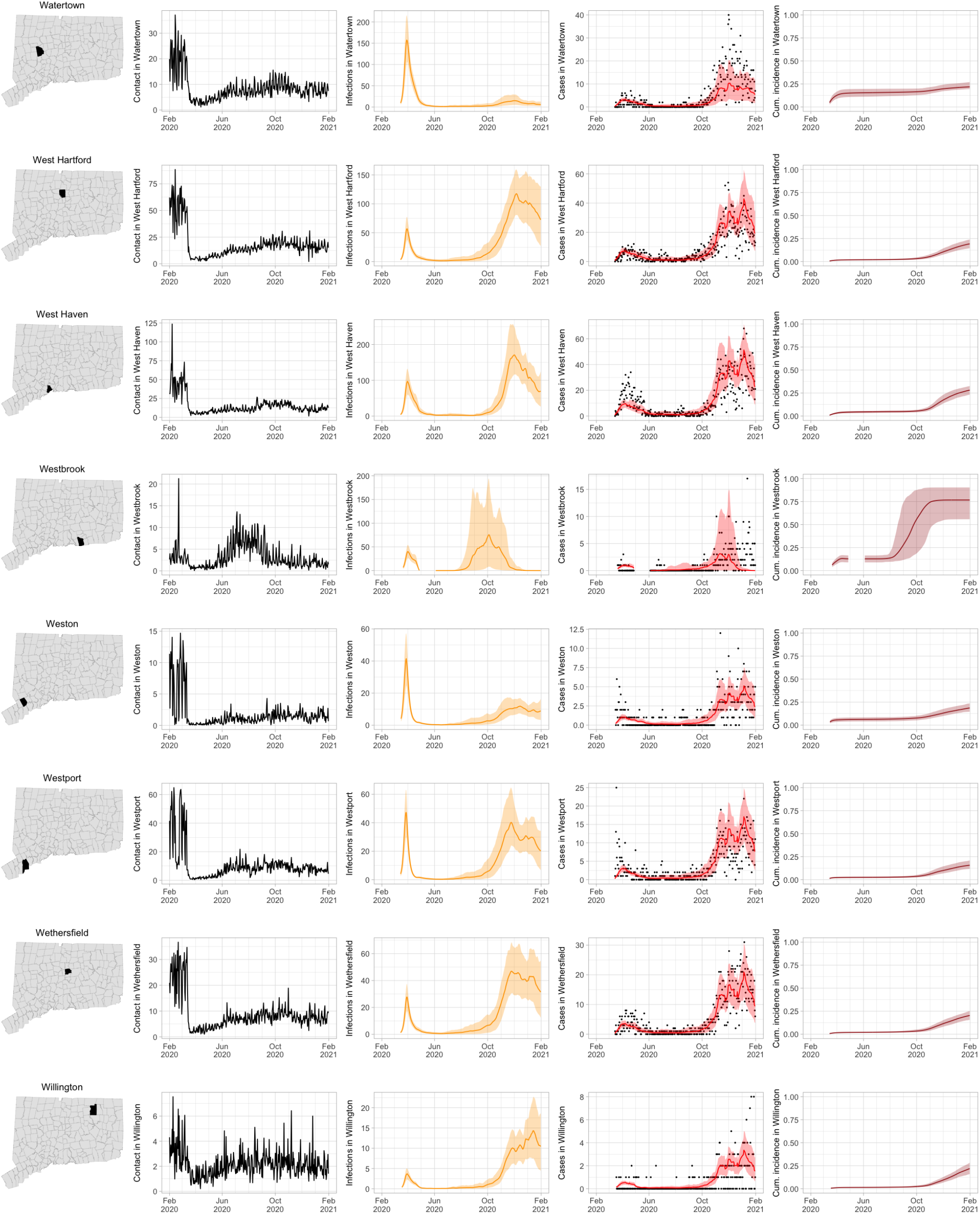
Contact for Connecticut towns and fitted SEIR model predictions with 95% uncertainty intervals.

**Figure 32:**
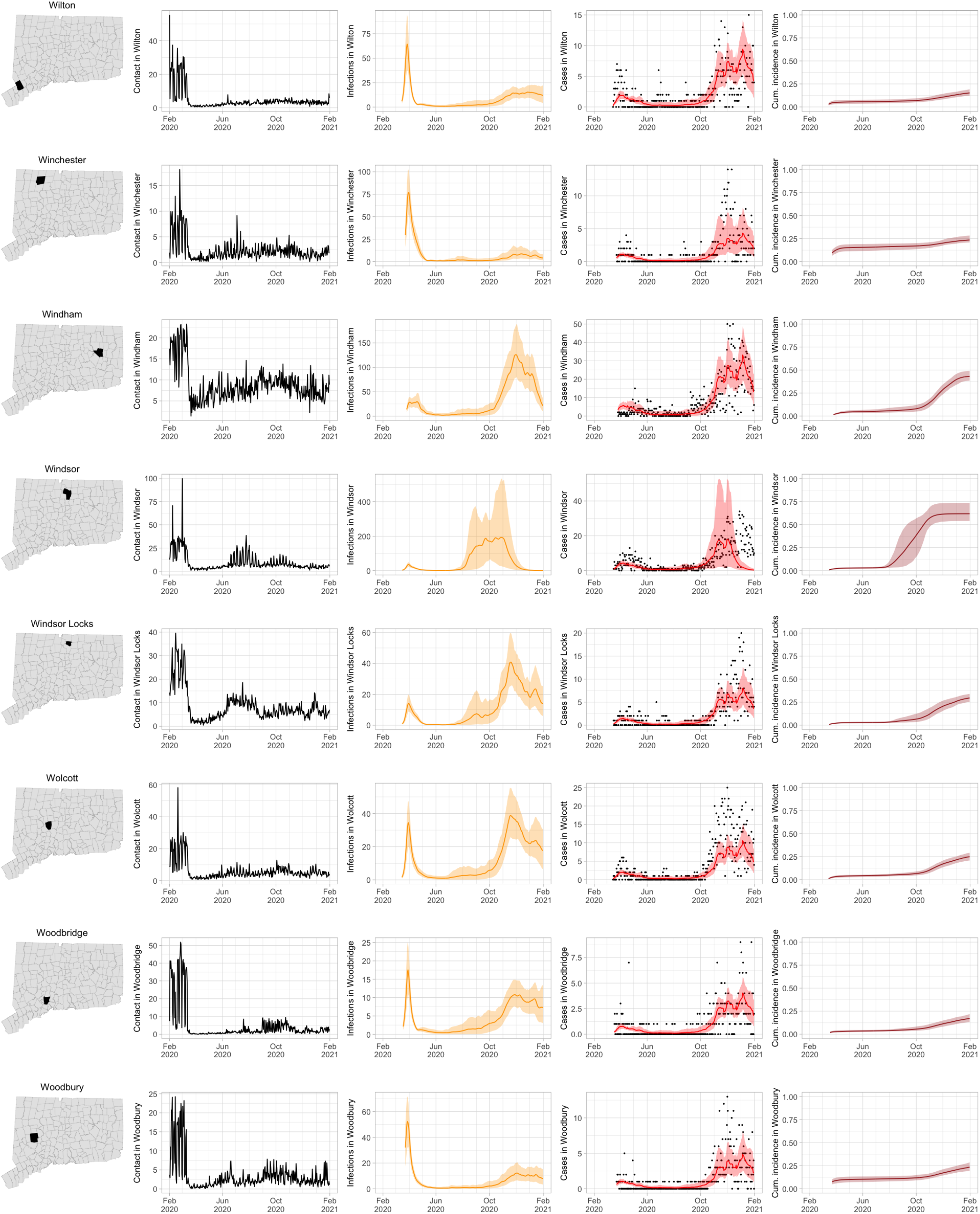
Contact for Connecticut towns and fitted SEIR model predictions with 95% uncertainty intervals.

**Figure 33:**
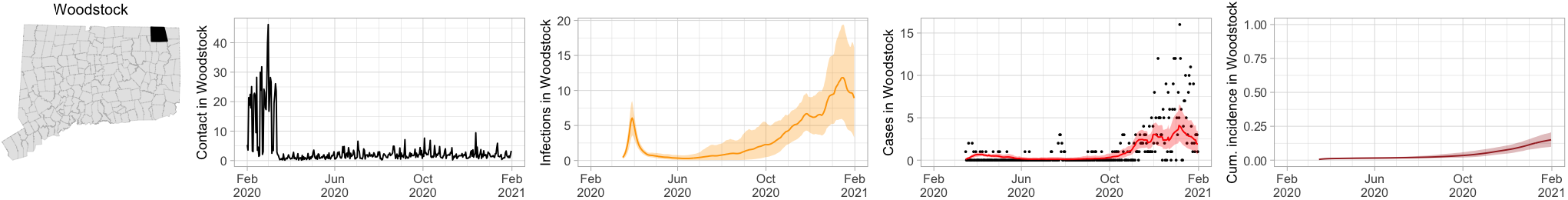
Contact for Connecticut towns and fitted SEIR model predictions with 95% uncertainty intervals.

## Footnotes

1 See e.g., 45 C.F.R. part 46, 21 C.F.R. part 56; 42 U.S.C. §241(d); 5 U.S.C. §552a; 44 U.S.C. §3501 et seq.

1 Aggregated mobility data were provided by Cuebiq, a location intelligence and measurement platform. Through its Data for Good program, Cuebiq provides access to aggregated mobility data for academic research and humanitarian initiatives. This first-party data is collected from anonymized users who have opted-in to provide access to their location data anonymously, through a GDPR-compliant framework. It is then aggregated to the census-block group level to provide insights on changes in human mobility over time.

